# County-Level Estimates of Excess Mortality Associated with COVID-19 in the United States

**DOI:** 10.1101/2021.04.23.21255564

**Authors:** Calvin A. Ackley, Dielle J. Lundberg, Lei Ma, Irma T. Elo, Samuel H. Preston, Andrew C. Stokes

**Author notes:** The authors would like to thank Robert N. Anderson and Farida B. Ahmad from the National Center for Health Statistics, Katherine Hempstead from the Robert Wood Johnson Foundation (RWJF), and Abe Dunn from the Bureau of Economic Analysis (BEA) for their input and technical support. Stokes gratefully acknowledges financial support from the RWJF. The views expressed in this paper are those of the authors and not necessarily the views of the BEA or RWJF.

## Abstract

**Background:** The COVID-19 pandemic in the U.S. has been largely monitored on the basis of death certificates containing reference to COVID-19. However, prior analyses reveal that a significant percentage of excess deaths associated with the pandemic were not directly assigned to COVID-19.

**Methods:** In the present study, we estimate a generalized linear model of expected mortality in 2020 based on historical trends in deaths by county of residence between 2011 and 2019. We use the results of the model to generate estimates of excess mortality and excess deaths not assigned to COVID-19 for 1,470 county-sets in the U.S. representing 3,138 counties.

**Results:** During 2020, more than one-fourth of U.S. residents (91.2 million) lived in counties where less than 75% of excess deaths were assigned to COVID-19. Across the country, we estimated that 439,698 excess deaths occurred in 2020, among which 86.7% were assigned to COVID-19. Some regions (Mideast, Great Lakes, New England, and Far West) reported the most excess deaths in large central metros, whereas other regions (Southwest, Southeast, Plains, and Rocky Mountains) reported the highest excess mortality in nonmetro areas. The proportion assigned to COVID-19 was lowest in large central metro areas (79.3%) compared to medium or small metros (87.4%), nonmetro areas (89.4%) and large fringe metros (95.2%). Regionally, the proportion of excess deaths assigned to COVID-19 was lowest in the Southeast (81.1%), Far West (81.2%), Southwest (82.6%), and Rocky Mountains (85.2%). Across the regions, the number of excess deaths exceeded the number of directly assigned COVID-19 deaths in the majority of counties. The exception to this was in New England, which reported more directly assigned COVID-19 deaths than excess deaths in large central metro areas, large fringe metros, and medium or small metros.

**Conclusions:** Across the U.S., many counties had substantial numbers of excess deaths that were not accounted for in direct COVID-19 death counts. Estimates of excess mortality at the local level can inform the allocation of resources to areas most impacted by the pandemic and contribute to positive protective behavior feedback loops (i.e. increases in mask-wearing and vaccine uptake).

## 1 Introduction

Estimates of excess deaths are critical to tracking the direct and indirect effects of the COVID-19 pandemic and for developing equitable policy responses.[1] Provisional estimates from the Center for Disease Control and Prevention (CDC) indicate that between 545,600 and 660,200 excess deaths occurred in the United States from January 26, 2020 to February 27, 2021.[2] The CDC further estimates that between 75 and 88% of excess deaths were directly assigned to COVID-19 on death certificates, suggesting that between 12 and 25% of excess deaths were not assigned to COVID-19.[2] Other prior estimates of excess mortality have also found significant discrepancies between direct COVID-19 deaths and excess mortality.[2, 3, 4, 5]

Excess deaths not assigned to COVID-19 may reflect a variety of factors, including COVID-19 deaths that were ascribed to other causes of death due to limited testing,[6] indirect deaths caused by interruptions in the provision of health care services,[7, 8] or indirect deaths caused by the broader social and economic consequences of the pandemic.[9, 10, 11] At the state-level, the percent of excess deaths not assigned to COVID-19 has been shown to vary significantly, suggesting that attribution of deaths to COVID-19 may not be uniform across the country.[4]

While prior estimates of excess mortality at the national and state levels are useful for understanding the impact of the pandemic on mortality levels broadly, estimation of excess mortality at the county-level may be valuable for several reasons. First, states are heterogeneous units in respect to sociodemographic characteristics, which contributes to geographic differences in mortality.[12, 13] Prior studies have found that the proportion of excess deaths not assigned to COVID-19 differed significantly by county-level sociodemographic and health care factors.[5, 14] Second, deaths are registered at the county-level.[15] Thus, it is reasonable to assume that administrative differences may exist between counties in the processing and assignment of deaths. Third, county-level data are essential for informing community and policy interventions. If a county’s direct COVID-19 tallies are substantially underestimated, measuring excess mortality at the county-level may be an important step to appreciating the full burden of the COVID-19 pandemic in an area and allocating response resources appropriately. Fourth, providing accurate data to residents could result in a positive behavior feedback loop, which encourages residents to understand the extent of mortality in their area and take protective actions such as wearing masks and pursuing vaccination.[16]

The objective of the present study is to generate estimates of excess mortality at the county-level and examine geographic variation in excess mortality and the proportion of excess deaths not assigned to COVID-19. Examining excess deaths at the county-level has the potential to identify counties with high excess mortality but low directly assigned COVID-19 mortality, indicating that COVID-19 is underreported on death certificates or substantial numbers of indirect deaths have occurred in these areas. These counties could represent regions that have been especially hard hit by the COVID-19 pandemic but whose mortality impacts did not appear in direct COVID-19 tallies and whose excess mortality has thus been hidden. In estimating excess mortality at the county-level, this study seeks to provide communities with estimates of the severity of the pandemic in their area which can be used to inform pandemic preparedness and response at the county, state, and national levels.

## 2 Data

We used provisional data from the National Center for Health Statistics (NCHS) on COVID-19 mortality and all-cause mortality by county of residence from January 1 to December 31, 2020 reported by June 3, 2021.^1^ We used data with a twenty-two week lag (December 31, 2020 to June 3, 2021) to improve the completeness of data, since prior analysis of provisional NCHS vital statistics reveal low completeness within the month following a death but more than 75 percent completeness after eight-weeks.[17] For counties with between 1 and 9 all-cause or COVID-19 deaths, NCHS censored the exact number of deaths, so we used the median value (5).

COVID-19 deaths were identified using the International Statistical Classification of Diseases and Related Health Problems, Tenth Revision (ICD-10) code *U* 07.1 and included deaths assigned to COVID-19 as the underlying cause as well as deaths in which COVID-19 was reported as a cause that contributed to death on the death certificate. Prior reports indicate that COVID-19 was assigned as the underlying cause on death certificate in 92% of deaths.[18]

For our historical comparison period, we used CDC Wonder data on all-cause mortality by county of residence from 2011 to 2019. For counties with fewer than 10 deaths, CDC Wonder censored the exact number of deaths, so we used a value of 5. To compute death rates, we used data on estimated populations from the U.S. Census Bureau for the years 2011 through 2020.

To assess geographic patterns in mortality, we classified counties into metropolitan-nonmetropolitan categories (large central metro, large fringe metro, small/medium metro, and nonmetro)[19]. The classifications were developed by the US Department of Agriculture (USDA) Economic Research Service (ERS) and modified by the National Center for Health Statistics. Large central metros included counties in metropolitan statistical areas with a population of more than 1 million. Large fringe metros were counties that surrounded the large central metros. Small or medium metros included counties in metropolitan statistical areas with a population between 50,000 and 999,999. Nonmetropolitan areas included all other counties. We also examined patterns by Bureau of Economic Analysis (BEA) Regions. These included New England (Connecticut, Maine, Massachusetts, New Hampshire, Rhode Island, and Vermont), Mideast (Delaware, District of Columbia, Maryland, New Jersey, New York, and Pennsylvania), Great Lakes (Illinois, Indiana, Michigan, Ohio, and Wisconsin), Plains (Iowa, Kansas, Minnesota, Missouri, Nebraska, North Dakota, and South Dakota); Southeast (Alabama, Arkansas, Florida, Georgia, Kentucky, Louisiana, Mississippi, North Carolina, South Carolina, Tennessee, Virginia, and West Virginia), Southwest (Arizona, New Mexico, Oklahoma, and Texas), Rocky Mountain (Colorado, Idaho, Montana, Utah, and Wyoming), and Far West (Alaska, California, Hawaii, Nevada, Oregon, and Washington). We then stratified these regions by metropolitan-nonmetropolitan categories to yield 32 distinct geographic units.

In order to provide precise estimates reflecting each county area, we aggregated small counties according to the United States Census Bureau’s County-Sets.^2^ Counties with a population of 50,000 or more generally stood alone, and smaller groups of counties were combined to form sets with a total population exceeding 50,000. Importantly, these county-sets were contiguous and preserved state borders. In using these county-sets, we were able to provide more precise estimates for geographies of sparsely populated counties than if we has modelled each of these small counties separately. Our data included 3138 counties with reported all-cause mortality for 2011-2020. This translated into a final analytic sample of 1,470 county sets.

The present investigation relied on de-identified publicly available data and was therefore exempted from review by the Boston University Medical Center Institutional Review Board. Analyses were conducted using R/R Studio. Additional details about the data along with programming code for replicating the analyses of the present study are available from the linked GitHub repository.^3^

## 3 Methodology

### 3.1 General Model for County-Level Mortality

To generate a prediction of expected mortality in 2020, we estimated a statistical model of mortality using historical mortality data from 2011-2019. Specifically, we modeled mortality at the county-set-year level using a quasi-poisson generalized linear model (QP-GLM) of the following form:^4,5^

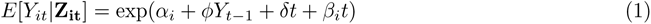

Here, *Y_it_* denoted the number of all-cause deaths divided by the total population of county *i* in year *t*. In the linear index, we included a county-set-specific intercept term, *α_i_*, which captured latent characteristics of each county-set that may be correlated with mortality. Importantly, this term picked up relevant information such as the distribution of age and health in each county-set.^6^ We included one lag of the dependent variable, *Y_t−_*_1_, to capture potential serial correlation in mortality.^7^ We included a time trend, *t*, and allowed the time trend to vary across county-sets according to *β_i_*. This accounts for the evolving distribution of age and other demographic characteristics across granular geographies that may be related to mortality.

We used the model estimates described above to compute fitted values of total deaths and the death rate per 100,000 person-years for each county-set and year from 2011-2020.^8^

### 3.2 Excess Deaths

We defined excess deaths as the difference between the number of predicted all-cause deaths in 2020 and the number of observed all-cause deaths in 2020. For each county in our sample, we produced an excess death rate for 2020 as well as a ratio of observed to expected deaths.

### 3.3 Excess Deaths Not Assigned to COVID-19

We defined excess deaths not assigned to COVID-19 as the difference between the number of excess deaths in 2020 and the number of observed directly assigned COVID-19 deaths in 2020. For each county in our sample, we decomposed the excess death rate into: (1) the observed death rate from COVID-19 and (2) the excess death rate not assigned to COVID-19. Next, we defined the proportion of excess deaths assigned to COVID-19 as the ratio of the direct COVID-19 death rate to the excess death rate.

### 3.4 Summary Statistics

We calculated summary statistics across all counties, metropolitan-nonmetropolitan categories, BEA regions, and BEA regions crossed with metropolitan-nonmetropolitan categories. In line with CDC guidance,[18] we excluded counties with negative excess death rates from the aggregations. In total, 48 county-sets representing less than 2% of the U.S. population (5.5 million residents) were excluded, yielding a sample size of 1,422 county-sets representing 3,066 counties.

## 4 Results

Across 1,422 county-sets in the U.S. representing 3,066 counties, we estimated that 439,698 excess deaths occurred in 2020. All-cause mortality was higher in most county-sets in 2020 compared to the years 2011 through 2019, leading to substantial excess mortality (Figure 1). The majority of counties also reported excess mortality not assigned to COVID-19. Across all counties, 86.7% of excess deaths were assigned to COVID-19, revealing that 13.3% of excess deaths were not assigned to COVID-19.

**Figure 1:**
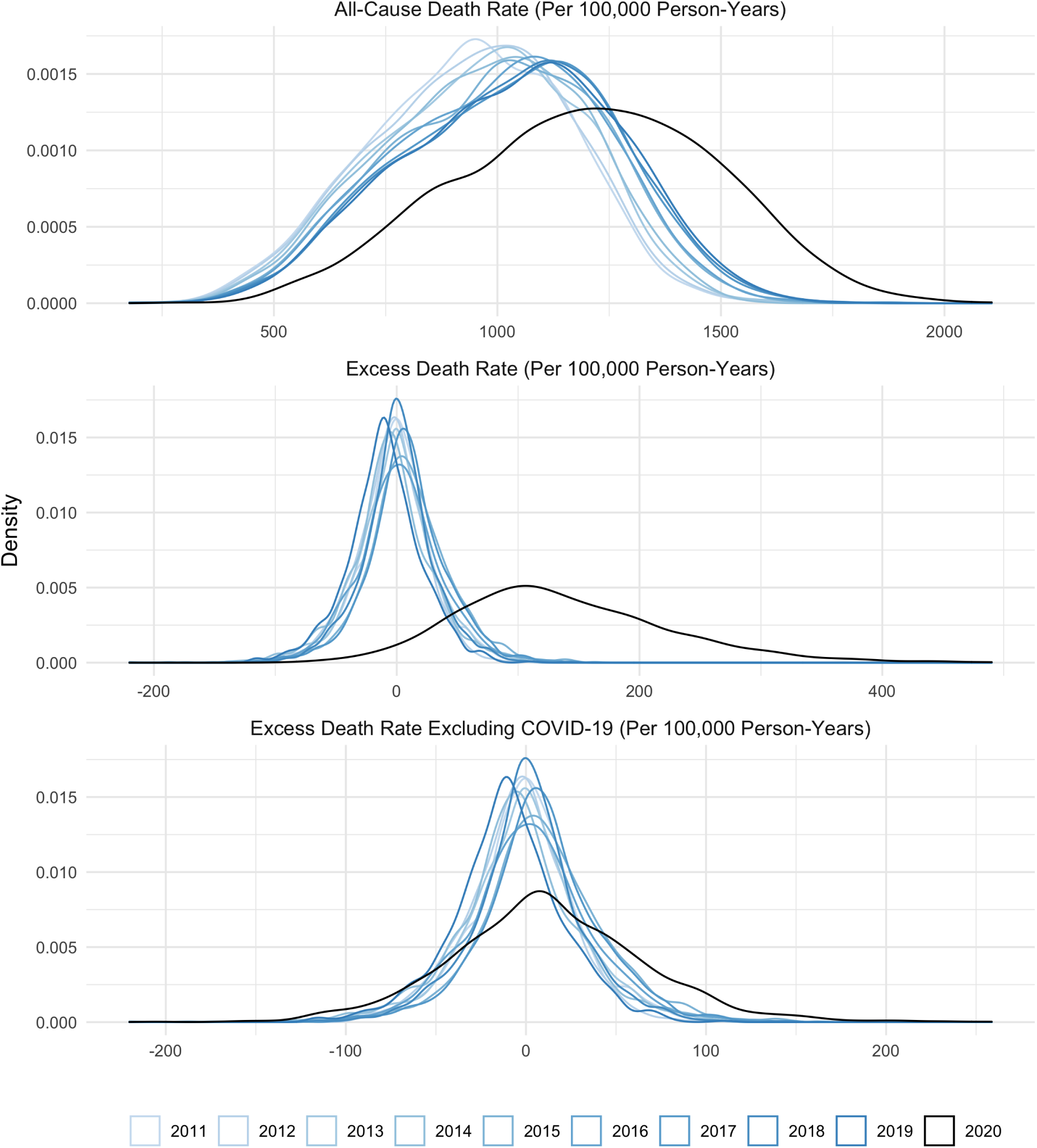
Distribution of All-Cause Deaths, Excess Deaths, and Excess Deaths Excluding COVID-19 Deaths per 100,000 Person-Years across U.S. Counties *Notes*: Expected deaths are computed as the in-sample fitted value for years 2011-2019. For 2020, expected deaths are computed as the out-of-sample fitted value.

**Table 1** presents summary statistics for excess mortality and excess deaths not assigned to COVID-19 across metropolitan-nonmetropolitan categories, BEA Regions, and metropolitan-nonmetropolitan categories crossed with BEA regions. Excess death rates were highest in nonmetro areas (159.9 deaths per 100,000 residents) and large metro areas (148.4 deaths per 100,000 residents) compared to small or medium metro areas (125.8 deaths per 100,000 residents) and large fringe metros (114.5 deaths per 100,000 residents). Across BEA regions, excess mortality was highest in the Mideast (191.5 deaths per 100,000 residents), Great Lakes (146.7 deaths per 100,000 residents), and Southwest (142.9 deaths per 100,000 regions) regions. **Figure 2** shows excess death rates in the U.S., highlighting counties with higher excess mortality in darker blue. This figure highlights the geographic dispersion of the COVID-19 pandemic with some areas reporting higher excess mortality than others, such as large central metros in the Mideast (279.9 deaths per 100,000 residents), nonmetro areas in the Southwest (206.0 deaths per 100,000 residents), nonmetro areas in the Southeast (184.5 deaths per 100,000 residents, and large central metros in the Great Lakes (181.7 deaths per 100,000 residents). The figure also identifies specific counties with the highest excess mortality such as Bronx and Queens, New York. **Appendix Table A1** presents summary statistics for excess mortality for each state, which further highlights the substantial geographic differences in excess mortality across the country.

**Figure 2:**
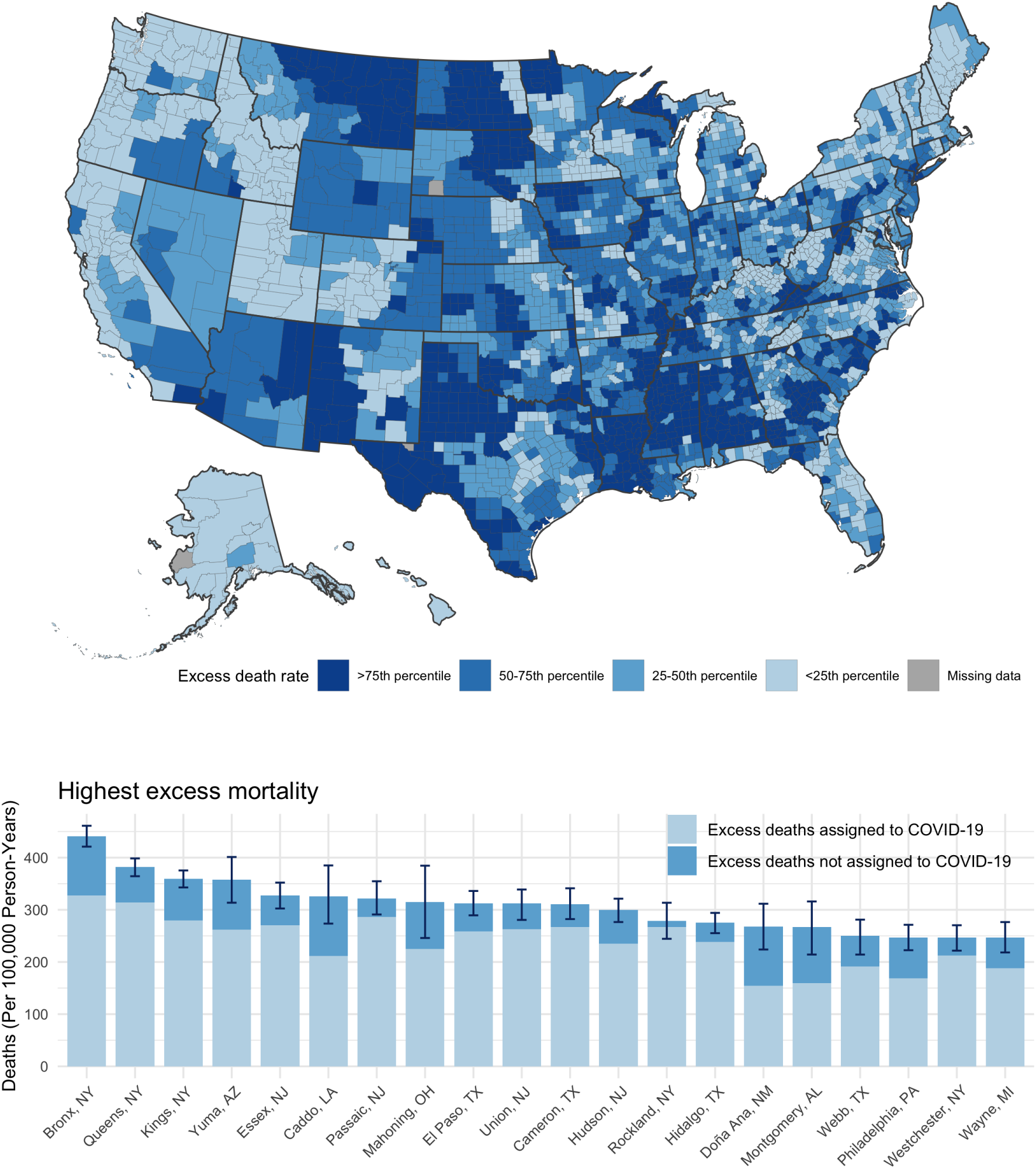
Excess Mortality Rates across U.S. Counties *Notes*: (top) U.S. counties labeled according to excess death rates, with the largest excess death rates in dark blue. (bottom) Counties with the highest excess mortality in the U.S. decomposed into directly assigned COVID-19 death rates and excess death rates not assigned to COVID-19. Limited to counties with populations of 200,000 residents or greater.

**Table 1:**
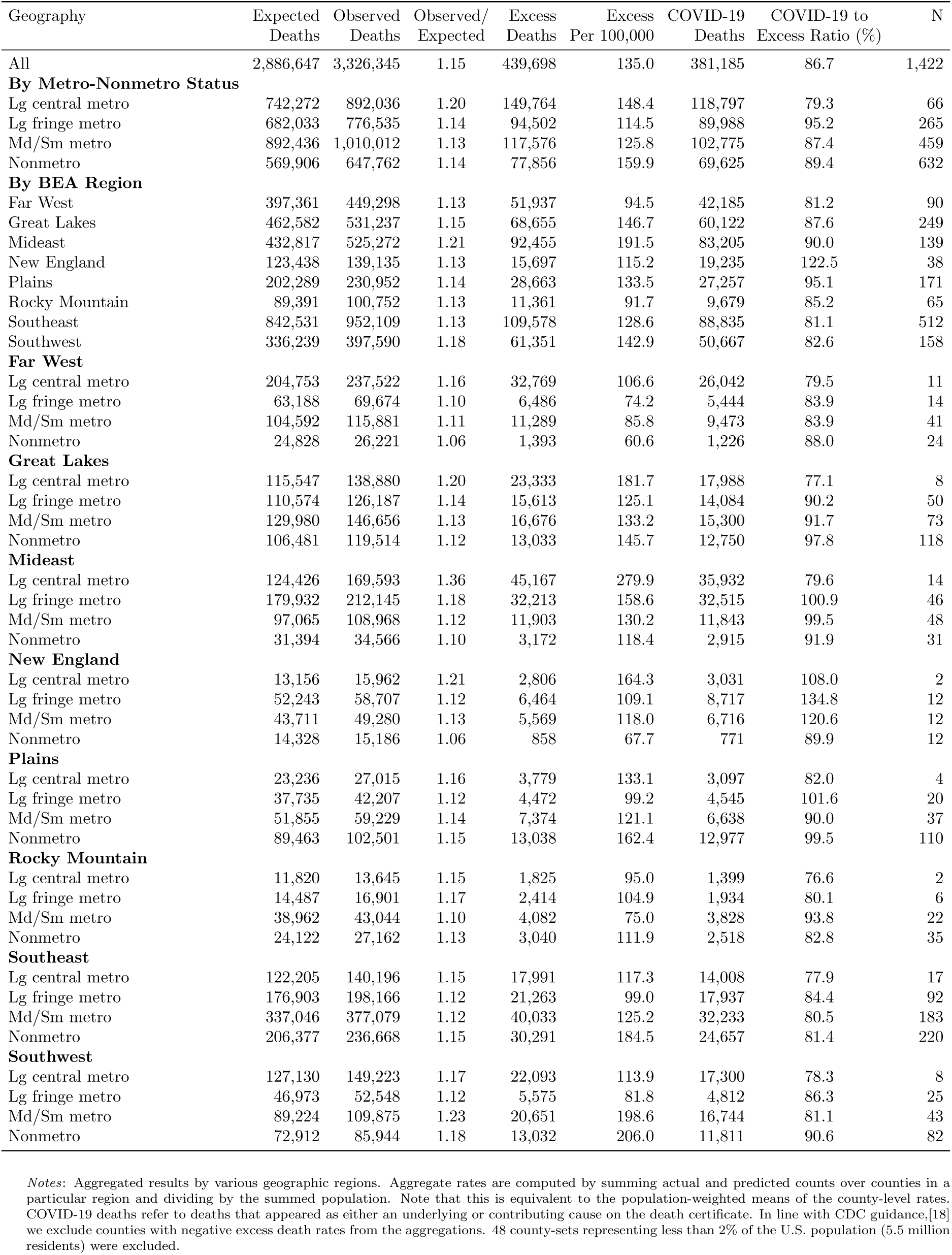
Excess Mortality and Direct COVID-19 Mortality by Metropolitan-Nonmetropolitan Status and BEA Region

Some regions (Mideast, Great Lakes, New England, and Far West) reported more excess deaths in large central metros compared to nonmetro areas. In contrast, the other regions (Southwest, Southeast, Plains, and Rocky Mountains) reported higher excess mortality in nonmetro areas compared to large central metros **(Appendix Figure A1)**. This difference demonstrates that patterns of excess mortality across metropolitan-nonmetropolitan categories varied by BEA region.

There was also significant variation in the proportion of excess deaths assigned to COVID-19 across counties in the U.S., with some counties reporting substantial numbers of excess deaths not assigned to COVID-19. Across the U.S., 91.2 million U.S. residents lived in counties where less than 75% of excess deaths were assigned to COVID-19 (**Appendix Table A2**). On average, assignment of excess deaths to COVID-19 was lowest in large central metro areas (79.3%) compared to medium or small metros (87.4%), nonmetro areas (89.4%) and large fringe metros (95.2%). Regionally, the proportion of excess deaths assigned to COVID-19 was lowest in the Southeast (81.1%), Far West (81.2%), Southwest (82.6%), and Rocky Mountains (85.2%).

**Figure 3** shows the percentage of excess deaths not assigned to COVID-19 across counties in the U.S., labeling counties as having high proportions of unassigned deaths (more than 25% excess deaths not assigned to COVID-19), moderate proportions of unassigned deaths (between 10 and 25% of excess deaths not assigned to COVID-19), or low proportions of unassigned deaths (between 0% and 10% of excess deaths not assigned to COVID-19).

**Figure 3:**
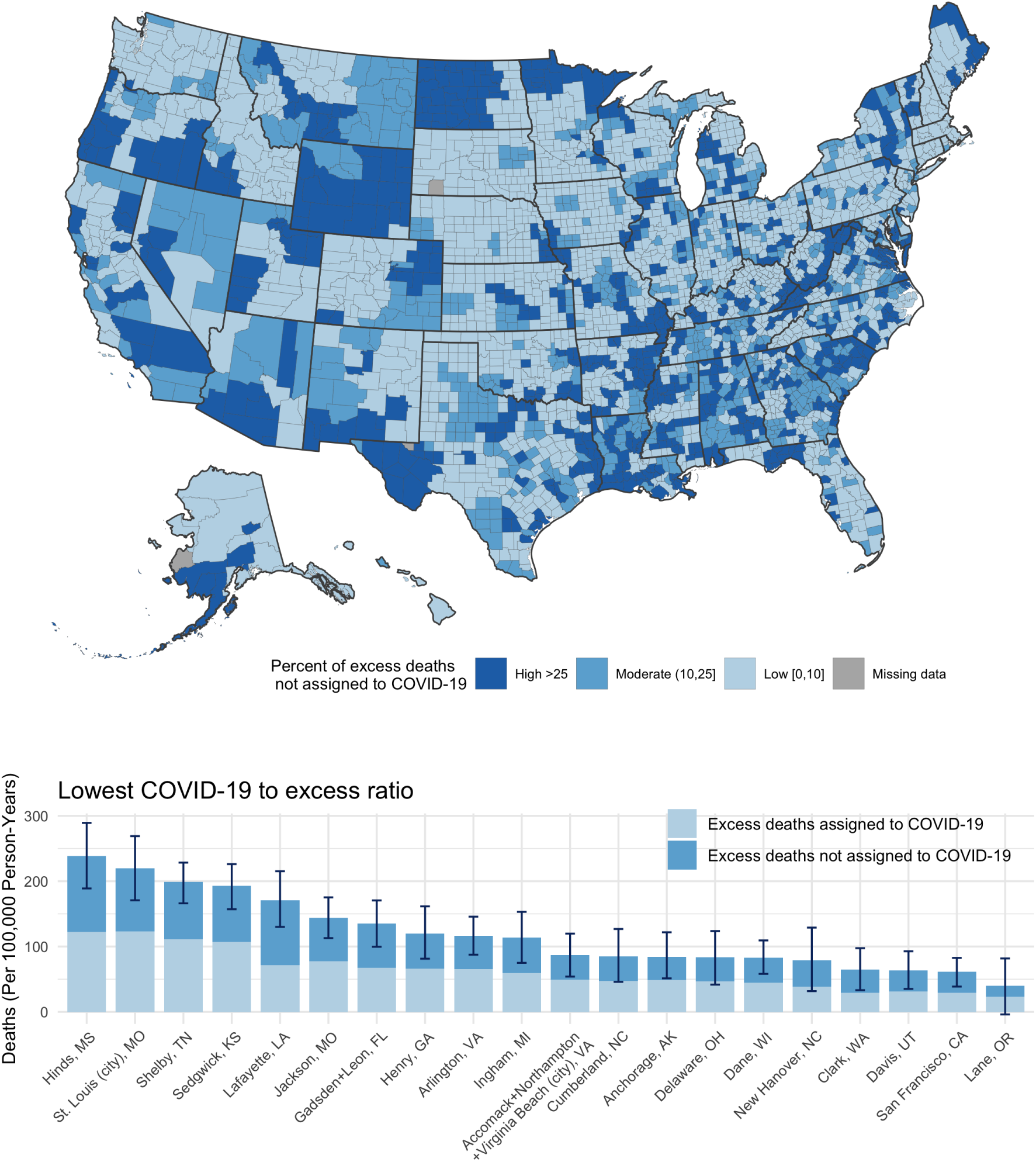
Proportion of Excess Deaths Not Assigned to COVID-19 across U.S. Counties *Notes*: (top) U.S. counties labeled according to the proportion of excess deaths not assigned to COVID-19, with the highest proportion of unassigned deaths in dark blue. (bottom) Counties with the highest proportion of unassigned deaths decomposed into directly assigned COVID-19 death rates and excess death rates not assigned to COVID-19. Limited to counties with populations of 200,000 residents or greater.

**Figure 4** plots excess death rates against directly assigned COVID-19 death rates for each county-set in each BEA region. In these plots, counties above the 45° line represent county-sets where excess deaths exceeded directly assigned COVID-19 deaths, meaning that excess deaths not assigned to COVID-19 occurred in these areas. All regions had some county-sets that reported excess deaths not assigned to COVID-19, and in all regions except New England, the majority of county-sets had excess deaths not assigned to COVID-19. In New England, directly assigned COVID-19 mortality exceeded excess mortality in most counties, indicating a negative excess death rate not assigned to COVID-19. Among these counties in New England, two types of counties emerged: (1) nonmetro counties with low direct COVID-19 death rates where excess death rates were negative, and (2) metro counties with positive excess death rates and substantial numbers of direct COVID-19 deaths that exceeded the number of excess deaths. **Appendix Figure A1** shows the proportion of excess deaths assigned to COVID-19 in New England across metropolitan-nonmetropolitan categories. Across large central metros, large fringe metros, and medium or small metros in New England, directly assigned COVID-19 deaths exceeded excess deaths. In nonmetro areas in New England, 67.7% of excess deaths were assigned to COVID-19, whereas 134.8% were assigned to COVID-19 in large fringe metro areas, 120.6% in medium or small metros, and 108.0% in large central metro areas.

**Figure 4:**
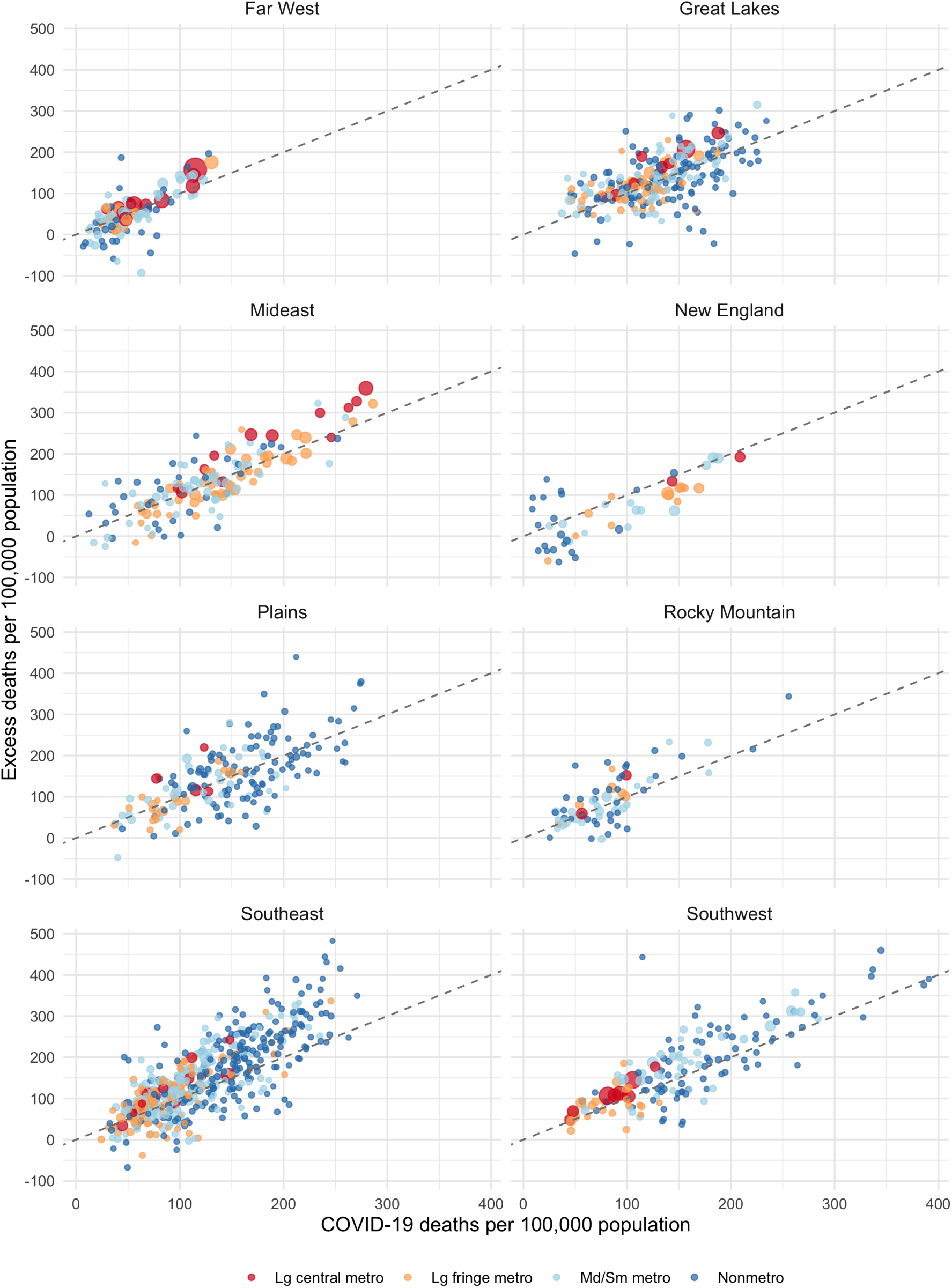
Plots of Excess Death Rates vs. Direct COVID-19 Death Rates by BEA Region *Notes*: Counties above the 45° line represent areas where excess deaths exceeded direct COVID-19 deaths, meaning the excess death rate not assigned to COVID-19 was positive. Counties below the 45° line represent areas where direct COVID-19 deaths exceeded excess deaths, meaning that the excess death rate not assigned to COVID-19 was negative. Excluded counties with direct COVID-19 death rates or excess death rates that were above the 99th percentile for each BEA region.

**Figure 5** decomposes excess death rates for county-sets with the highest excess death rates not assigned to COVID-19 and the most negative excess death rates not assigned to COVID-19. Most counties across the U.S. experienced substantial numbers of excess deaths not assigned to COVID-19, and many of these excess deaths not assigned to COVID-19 were significantly positive. Counties with large, negative excess mortality not assigned to COVID-19 were mostly in the New England region such as Barnstable/Dukes/Nantucket, Massachusetts, Essex, Massachusetts, Hillsborough, New Hampshire, Bristol/Providence, Rhode Island, and Middlesex, Massachusetts. Other counties outside of New England with statistically significant negative excess death rates not assigned to COVID-19 included Brevard, Florida and Hawaii County, Hawaii.

**Figure 5:**
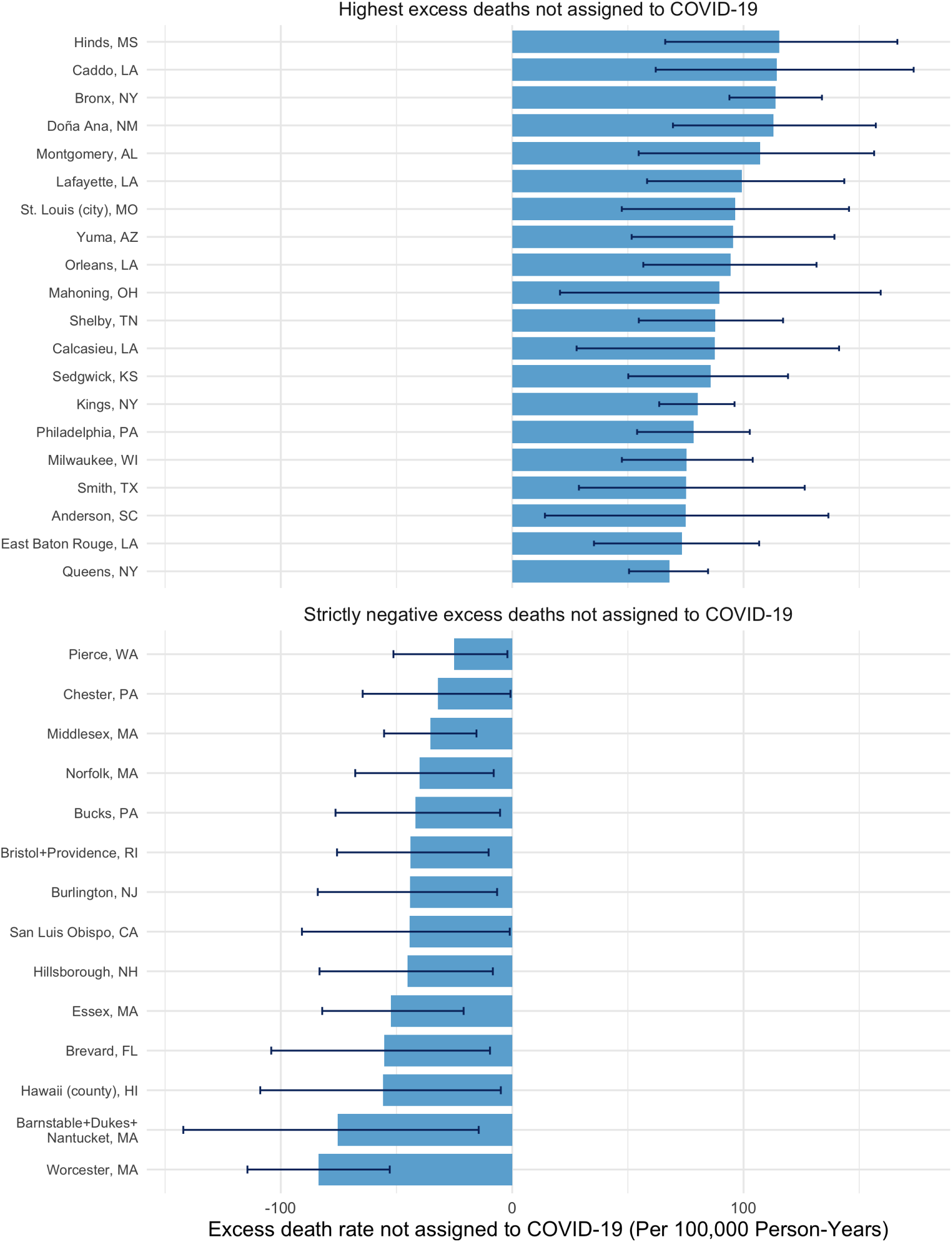
U.S. Counties with the Highest Excess Death Rates Not Assigned to COVID-19 and Statistically Significant Negative Excess Death Rates Not Assigned to COVID-19 *Notes*: (top) U.S. county-sets with the highest, statistically significant positive excess death rates not assigned to COVID-19. (bottom) U.S. county-sets with statistically significant negative excess death rates not assigned to COVID-19. The 95% confidence intervals indicate uncertainty in the excess death estimates as calculated by the bootstrapping procedure. Limited to counties with populations of 200,000 residents or greater.

**Appendix Figure A2** and **Appendix Figure A3** display a time series of observed death rates from 2011 to 2019 and a comparison of the 2020 predicted death rate and the 2020 expected death rate. In the East South Central region where a relatively large share of excess deaths were not assigned to COVID-19, the observed death rate in 2020 substantially exceeded the expected 2020 death rate and the expected 2020 death rate plus COVID-19 deaths. In New England, however, where direct COVID-19 deaths exceeded excess deaths, the opposite occurred – with the expected death rate plus COVID-19 deaths exceeding the observed death rate in 2020.

Only 48 county-sets, representing less than 2% of the U.S. population (5.5 million residents) experienced negative excess mortality (where observed mortality was less than expected mortality). Among these counties, no county-sets had statistically significant negative excess death rates where the uncertainty intervals for the excess death rates did not overlap with 0. This suggests that most counties across the U.S. experienced positive excess mortality in 2020. **Appendix Table C1** provides estimates of excess death rates and their uncertainty intervals for 1,470 county-sets across the U.S..

## 5 Discussion

In this study, we produced county-level estimates of excess mortality and the proportion of excess deaths not assigned to COVID-19 and examined geographic variation in mortality across the United States. We found that more than one-forth of U.S. residents (91.2 million) lived in counties where less than 75% of excess deaths were assigned to COVID-19, suggesting that the impact of the COVID-19 pandemic has not been fully registered in many parts of the country. Assignment of excess deaths to COVID-19 was particularly low in the Southeast, Far West, Rocky Mountain, and Southwest regions and in large central metro areas compared to other areas. New England was unique in reporting more directly assigned COVID-19 deaths than excess deaths.

Across counties in the U.S., we estimated that 439,698 total excess deaths occurred in 2020, of which 86.7% were assigned to COVID-19. This estimate is similar to an estimate of 458,000 excess deaths in the during 2020 produced by Islam et al.[21] Recent estimates by Woolf et al. calculated 522,368 excess deaths between March 1, 2020, and January 2, 2021, which is higher than our estimate.[4] Our estimate may be more conservative than Woolf et al. because we incorporated historical data from 2011 through 2019 for our calculations of expected mortality whereas Woolf et al. used trends from 2014 through 2019. Using data from 1999 to 2020 and a cubic model, Glei identified 402,743 excess deaths in 2020, which was lower than our estimate.[22] Ahmad et al. found an increase of 503,976 deaths between 2019 and 2020, which is also higher than our estimate of excess mortality.[23] However, this estimate did not control for recent mortality trends.

Our study also reveals substantial heterogeneity in excess deaths and the proportion of excess deaths not assigned to COVID-19 across counties, which are the administrative unit for death registration. This highlights the value of studying excess mortality at the county-level since state and national-estimates mask significant variability within states. This finding is in line with studies of excess deaths in California that found significant differences in excess deaths by race/ethnicity and education, which vary by county.[24, 25]

Consistent with previous results, we find evidence of wide gaps between excess mortality and directly assigned COVID-19 mortality in many areas across the United States. There are several potential explanations for the discrepancy between excess mortality and directly assigned COVID-19 mortality. One explanation is that the gap reflects underreporting of COVID-19 deaths. Especially early in the pandemic, testing was severely limited, which may have reduced the likelihood of COVID-19 being assigned to the death certificate. Underreporting may have also related to a lack of awareness of the clinical manifestations of COVID-19 early in the pandemic as well as various social, health care, and political factors.[26, 5, 14] In addition to underreporting, gaps between excess and direct mortality may in part be explained by the indirect effects of the pandemic on mortality levels. Indirect effects could relate to interruptions or delays in health care or the broader social and economic upheaval caused by the pandemic, including loss of employment, social isolation and loneliness, and other factors.[27, 28, 29] Many states have reported increases in overdose deaths during the COVID-19 pandemic, and NCHS data suggest that approximately 19,000 more deaths from unintentional injuries occurred in 2020 than in 2019.[23]

Our study made use of data through December 31, 2020 reported by June 3, 2021, meaning that deaths could be reported for up to five months after they occurred. As a result, our estimate of the proportion of excess deaths not assigned to COVID-19 should be interpreted accordingly, indicating that after accounting for five months of potential reporting and processing delays, 13.3% of excess deaths were still not assigned to COVID-19. It is important to acknowledge that COVID-19 mortality surveillance data sources such as local health system dashboards that the public uses to interact with county-level data in real-time more substantially underestimate excess mortality due to significant delays in COVID-19 reporting that are not accounted for. Similarly, our estimate of the proportion of excess deaths not assigned to COVID-19 (13.3%) is lower than a previous study using county-level data (17%), which only incorporated a two and a half month delay between occurrence and reporting.[5]

We also observed a number of counties in which the direct COVID-19 death rate exceeded our estimates of excess mortality, especially in New England. These included two types of counties: (1) nonmetro areas that experienced few direct COVID-19 deaths but negative excess mortality and (2) metro areas that had positive excess mortality but where direct COVID-19 deaths exceeded excess deaths. A similar pattern was observed internationally, with countries such as New Zealand and Taiwan having few direct COVID-19 deaths and negative excess mortality while countries such as Luxembourg, France, Belgium, and Costa Rica had positive excess mortality but their direct COVID-19 death rates exceeded their excess death rates.[30] This finding could have occurred for several reasons. First, increases in mortality in 2020 due to COVID-19 may have been offset by declines in deaths from other causes. Provisional data on cause of death for 2020 indicates that flu deaths declined significantly relative to prior years, which may explain part of the offset.[31] Other causes of death may also have experienced reductions in 2020. For example NCHS data indicates that there were approximately 2,600 fewer suicide deaths in 2020 relative to 2019.[23] Shelter-in-place policies may have also been associated with reductions in non-natural deaths.[32] While we did not examine associations with income and education in this study, several of the counties (Barnstable/Dukes/Nantucket, Massachusetts, Bristol/Providence, Rhode Island, and Middlesex, Massachusetts) stood out as economically privileged areas that may have been isolated from the impacts of the pandemic through an ability to work-from-home and avoid household crowding. Another reason direct Covid-19 deaths may have exceeded our estimates of excess deaths is if medical certifiers in a county over-assigned COVID-19 to death certificates. In addition, the differences could at least partially relate to how directly assigned COVID-19 deaths were counted by NCHS; while COVID-19 was listed as the underlying cause in the vast majority of cases, in about 8% of cases, COVID-19 was listed as a contributing cause and still directly assigned to COVID-19. Finally, frailty selection may have occurred if deaths from COVID-19 occurred among individuals who were likely to die from other causes, resulting in reductions in those causes of death. It is also important to note that in instances where directly assigned COVID-19 deaths exceeded excess deaths in the overall population, patterns could differ among population subgroups related to age, race/ethnicity, and income.

Accurate county-level predictions of excess mortality, and their associated levels of uncertainty, are an important addition to existing work on excess mortality at the state and national level. County-level estimates can enable researchers to examine geographic variation in excess mortality and leverage county-level variation in sociodemographic, health, and structural factors to examine inequities in excess deaths. They may also be relevant to local public health departments, to be used in conjunction with their existing efforts to monitor deaths directly assigned to COVID-19. Accurate death tallies at the county-level may play an important role in motivating individual and community responses, including vaccine uptake. Our study indicates that directly assigned COVID-19 death rates have been less accurate measures of excess mortality in areas such as the Southeast and Rocky Mountain regions, which are also areas that are experiencing the slowest vaccine uptake.[33]

This analysis had several limitations. First, unlike prior state-level analyses which leveraged weekly data on deaths, the present study used cumulative data on COVID-19 and all-cause mortality for all of 2020. Given this limitation in the available data, it was not possible to examine changes in excess mortality over time or trends in the proportion of deaths not assigned to COVID-19 at the county level. Examining trends in excess mortality using small-area data is a priority for future research which may help to distinguish the direct effects of the pandemic from indirect consequences associated with interruptions in health care and the social and economic consequences of pandemic response measures. Second, when considering patterns of excess mortality across the United States, an important caveat is that age structure differs across counties. Since COVID-19 mortality is more common in older populations, some of the patterns observed across counties may simply reflect differences in age structure. Thus, an important future direction for county-level analyses of excess mortality is to age standardize the estimates when age-specific mortality data become available. Third, the provisional county-level mortality files released by the NCHS did not include information on cause of death, and therefore it was not possible to disentangle the sources of excess deaths in 2020. Decomposing excess deaths by cause of death will be critical to understanding why some counties have a higher fraction of unassigned deaths than others and the extent to which the discrepancies are explained by COVID-19 death undercounts versus indirect pandemic effects. For example, such an analysis might partition natural from non-natural deaths under the assumption that non-natural causes of death are unlikely to represent misasacribed COVID-19 deaths. Information on cause of death will also be valuable for understanding the extent to which declines in mortality from other causes have offset COVID-19 deaths, thereby leading to smaller estimates of both excess deaths and the percent of excess deaths that were not assigned to COVID-19. Given the potential for offset from other causes of death, it is likely our overall finding that 13.3% of excess deaths were not assigned to COVID-19 represents a lower bound on the percent unassigned. Finally, the data used in the present study are provisional in nature and may be subject to further corrections by the NCHS in the process of generating final death counts by cause of death for 2020.

In conclusion, the present study builds on prior work by extending estimates of excess mortality and excess deaths not assigned to COVID-19 to US counties. The added geographic detail of these estimates compared to prior studies may facilitate research on the causes and consequences of the COVID-19 pandemic on population health and provide useful data for local area health policy and planning. Estimates of excess mortality at the local level can also inform the allocation of resources to areas most impacted by the pandemic and contribute to positive protective behavior feedback loops (i.e. increases in mask-wearing and vaccine uptake). In doing so, they can inform the response to the COVID-19 pandemic and to any future pandemics that the country may face.

## Data Availability

The present investigation used publicly available data from the National Center for Health Statistics and the U.S. Census Bureau.

https://data.cdc.gov/NCHS/AH-County-of-Residence-COVID-19-Deaths-Counts-2020/75vb-d79q

https://github.com/pophealthdeterminantslab/county-level-estimates-of-excess-mortality

## 6 Appendix A: Tables and Figures

**Figure A1:**
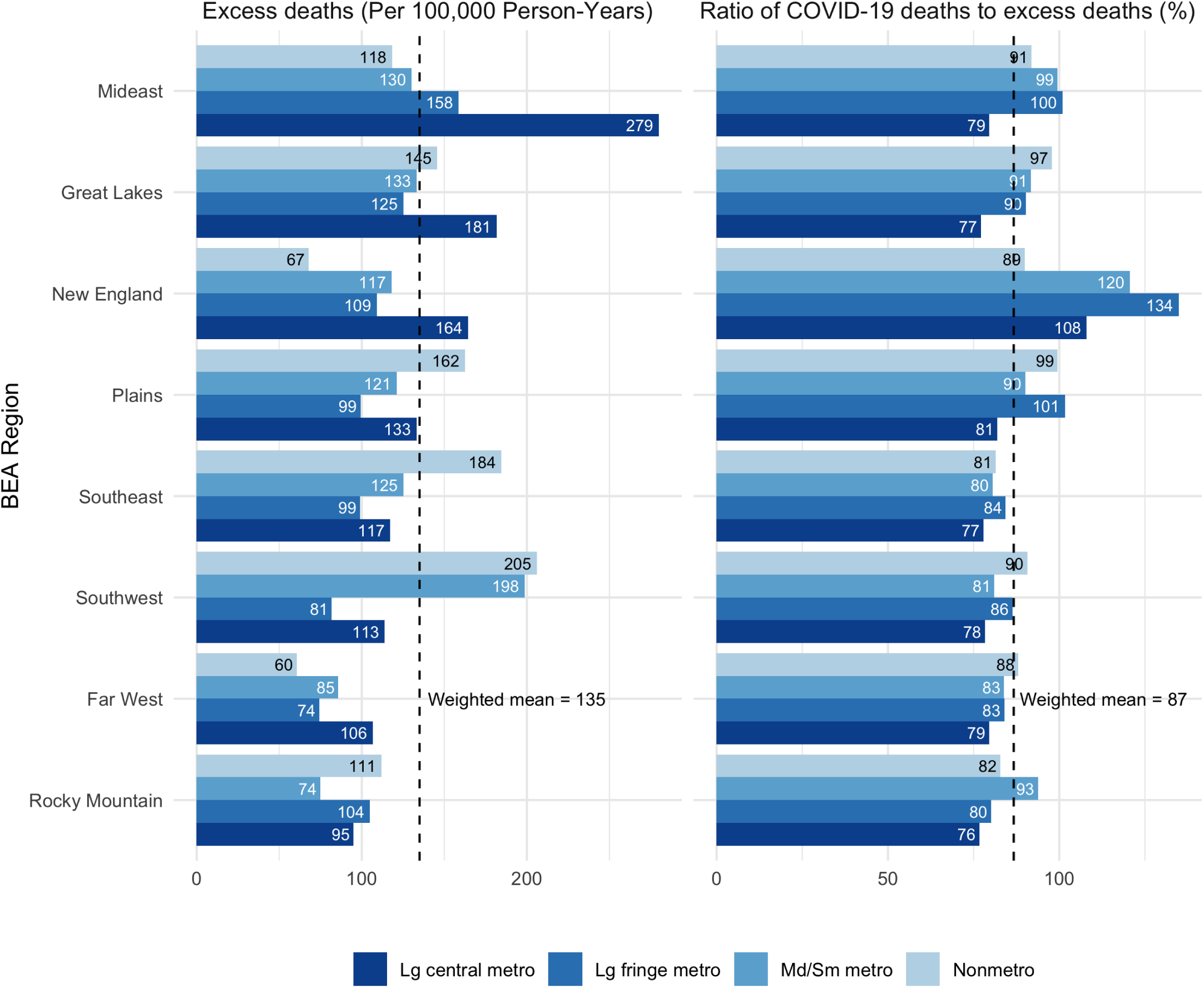
Excess Death Rates and Proportion of Excess Deaths Assigned to COVID-19 by BEA Region and Metropolitan-Nonmetropolitan Status *Notes*: Aggregated results by various geographic regions. Aggregate rates are computed by summing actual and predicted counts over counties in a particular region and dividing by the summed population. Note that this is equivalent to the population-weighted means of the county-level rates. COVID-19 deaths refer to deaths that appeared as either an underlying or contributing cause on the death certificate. In line with CDC guidance,[18] 48 county-sets with negative excess mortality representing less than 2% of the U.S. population (5.5 million residents) were excluded.

**Figure A2:**
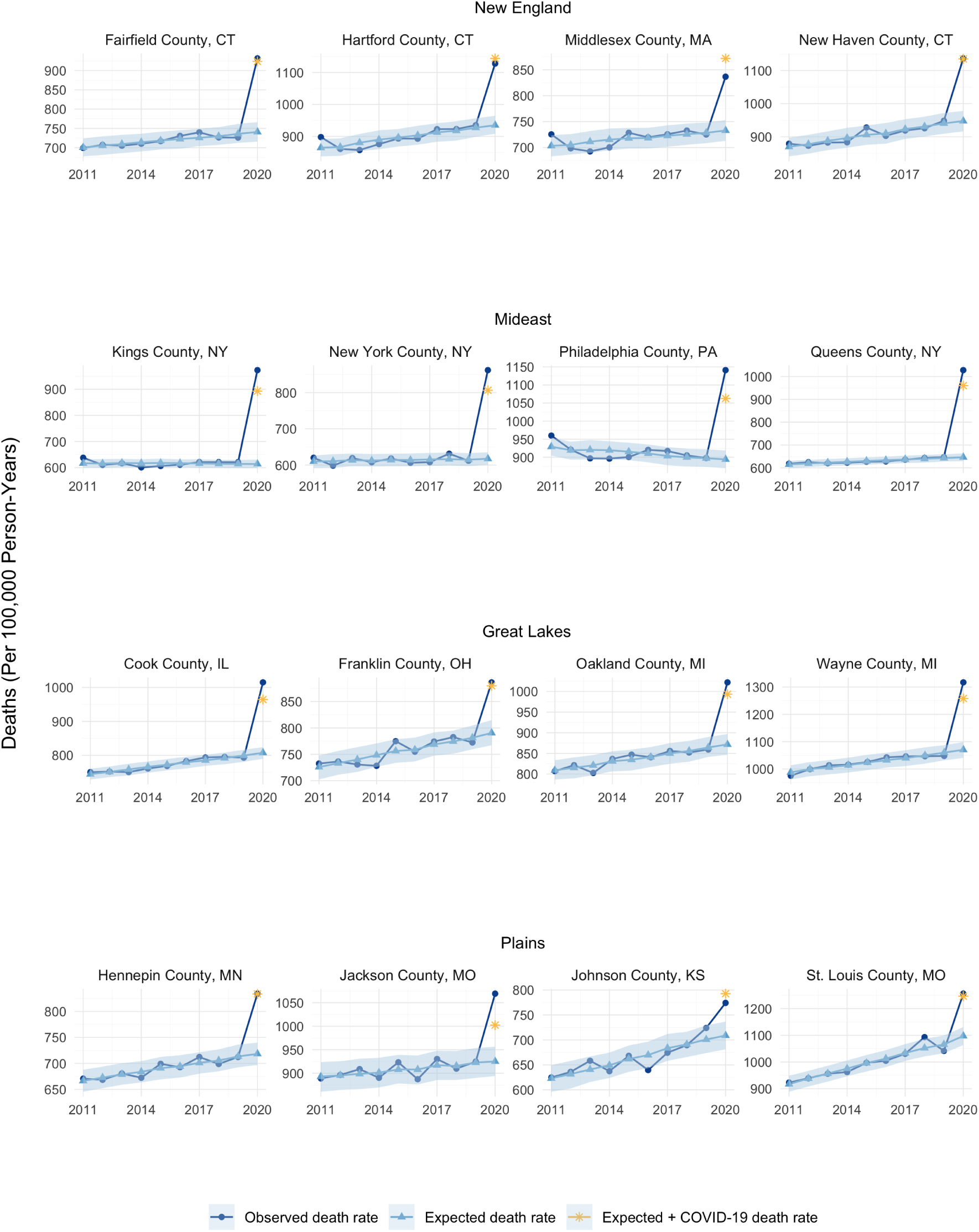
Comparison of Observed and Expected Deaths in Large Counties, 2011-2020 *Notes*: Observed and expected deaths for largest counties by population in each BEA Region from 2011-2020. Expected death counts are generated using a Poisson GLM with data from 2011-2019. The shaded area represents a 95% prediction region for each point. These regions are generated using a parametric bootstrap.

**Figure A3:**
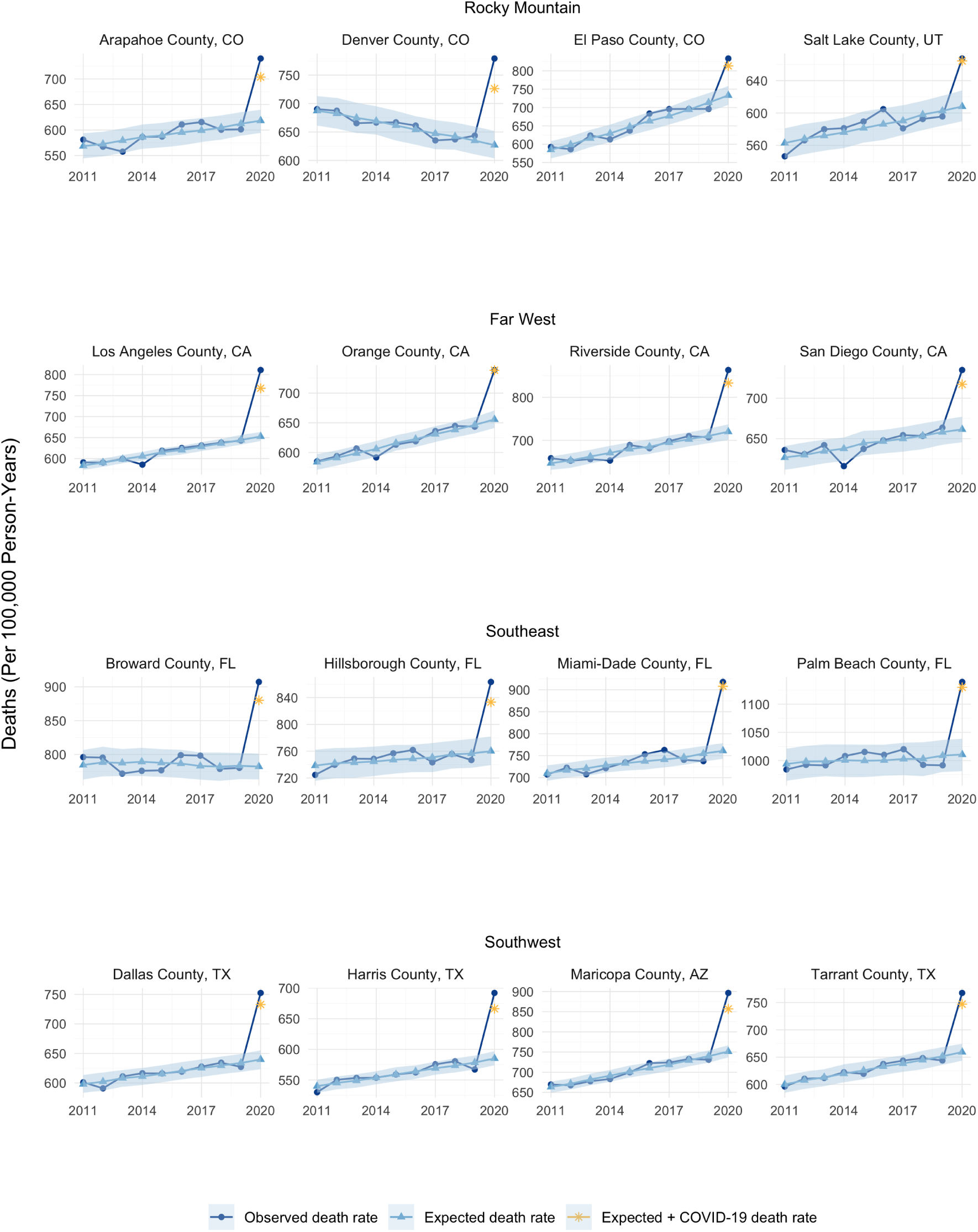
Comparison of Observed and Expected Deaths in Large Counties, 2011-2020 *Notes*: Observed and expected deaths for largest counties by population in each BEA Region from 2011-2020. Expected death counts are generated using a Poisson GLM with data from 2011-2019. The shaded area represents a 95% prediction region for each point. These regions are generated using a parametric bootstrap.

**Table A1:** Excess Mortality and COVID-19 Mortality by State

| State | Expected Deaths | Observed Deaths | Observed/Expected | Excess Deaths | Excess Per 100,000 | COVID-19 Deaths | COVID-19 to Excess Ratio (%) | N |
| --- | --- | --- | --- | --- | --- | --- | --- | --- |
| AK | 4,181 | 4,630 | 1.11 | 449 | 67.4 | 269 | 59.9 | 6 |
| AL | 55,639 | 64,776 | 1.16 | 9,137 | 186.0 | 6,991 | 76.5 | 40 |
| AR | 33,610 | 38,099 | 1.13 | 4,489 | 147.9 | 4,014 | 89.4 | 32 |
| AZ | 63,304 | 75,741 | 1.20 | 12,437 | 167.8 | 9,182 | 73.8 | 11 |
| CA | 269,553 | 312,029 | 1.16 | 42,476 | 108.7 | 33,004 | 77.7 | 41 |
| CO | 40,525 | 46,516 | 1.15 | 5,991 | 103.4 | 4,926 | 82.2 | 23 |
| CT | 31,986 | 37,831 | 1.18 | 5,845 | 164.0 | 6,214 | 106.3 | 8 |
| DC | 5,035 | 6,198 | 1.23 | 1,163 | 162.5 | 885 | 76.1 | 1 |
| DE | 9,868 | 11,071 | 1.12 | 1,203 | 121.8 | 1,038 | 86.3 | 3 |
| FL | 214,336 | 237,542 | 1.11 | 23,206 | 106.7 | 21,324 | 91.9 | 48 |
| GA | 89,034 | 102,400 | 1.15 | 13,366 | 125.4 | 10,257 | 76.7 | 67 |
| HI | 9,204 | 9,580 | 1.04 | 376 | 33.1 | 321 | 85.3 | 2 |
| IA | 30,695 | 35,413 | 1.15 | 4,718 | 153.1 | 4,725 | 100.2 | 31 |
| ID | 13,512 | 14,740 | 1.09 | 1,228 | 74.1 | 1,393 | 113.5 | 13 |
| IL | 111,763 | 132,652 | 1.19 | 20,889 | 165.1 | 17,268 | 82.7 | 51 |
| IN | 68,213 | 78,017 | 1.14 | 9,804 | 145.2 | 9,337 | 95.2 | 50 |
| KS | 28,040 | 32,045 | 1.14 | 4,005 | 137.5 | 3,588 | 89.6 | 25 |
| KY | 50,114 | 55,041 | 1.10 | 4,927 | 110.9 | 4,518 | 91.7 | 44 |
| LA | 46,735 | 56,741 | 1.21 | 10,006 | 215.7 | 7,081 | 70.8 | 35 |
| MA | 60,873 | 68,213 | 1.12 | 7,340 | 105.3 | 10,103 | 137.6 | 12 |
| MD | 52,411 | 60,073 | 1.15 | 7,662 | 126.1 | 6,706 | 87.5 | 19 |
| ME | 9,162 | 9,527 | 1.04 | 365 | 45.1 | 256 | 70.2 | 6 |
| MI | 100,676 | 115,750 | 1.15 | 15,074 | 152.0 | 12,382 | 82.1 | 49 |
| MN | 46,670 | 52,370 | 1.12 | 5,700 | 100.0 | 5,932 | 104.1 | 34 |
| MO | 64,756 | 73,887 | 1.14 | 9,131 | 148.2 | 7,774 | 85.1 | 45 |
| MS | 33,562 | 40,176 | 1.20 | 6,614 | 221.9 | 5,023 | 75.9 | 36 |
| MT | 10,594 | 12,022 | 1.13 | 1,428 | 132.1 | 1,274 | 89.2 | 11 |
| NC | 97,101 | 106,818 | 1.10 | 9,717 | 93.3 | 8,540 | 87.9 | 61 |
| ND | 6,497 | 8,054 | 1.24 | 1,557 | 202.0 | 1,337 | 85.8 | 8 |
| NE | 17,395 | 19,540 | 1.12 | 2,145 | 109.8 | 2,306 | 107.5 | 18 |
| NH | 6,489 | 6,928 | 1.07 | 439 | 60.0 | 652 | 148.5 | 2 |
| NJ | 76,860 | 95,965 | 1.25 | 19,105 | 213.6 | 17,927 | 93.8 | 21 |
| NM | 19,943 | 23,749 | 1.19 | 3,806 | 181.3 | 3,012 | 79.1 | 17 |
| NV | 26,962 | 30,617 | 1.14 | 3,655 | 115.6 | 3,462 | 94.7 | 5 |
| NY | 151,125 | 196,520 | 1.30 | 45,395 | 242.5 | 38,404 | 84.6 | 45 |
| OH | 127,267 | 142,815 | 1.12 | 15,548 | 133.2 | 14,930 | 96.0 | 61 |
| OK | 41,600 | 47,795 | 1.15 | 6,195 | 156.3 | 5,548 | 89.6 | 33 |
| OR | 36,631 | 38,587 | 1.05 | 1,956 | 48.0 | 1,626 | 83.1 | 18 |
| PA | 137,517 | 155,445 | 1.13 | 17,928 | 139.8 | 18,245 | 101.8 | 50 |
| RI | 10,342 | 11,819 | 1.14 | 1,477 | 139.5 | 1,869 | 126.5 | 4 |
| SC | 53,223 | 61,179 | 1.15 | 7,956 | 152.6 | 5,782 | 72.7 | 30 |
| SD | 8,236 | 9,643 | 1.17 | 1,407 | 158.8 | 1,595 | 113.4 | 10 |
| TN | 74,695 | 84,197 | 1.13 | 9,502 | 137.8 | 7,452 | 78.4 | 46 |
| TX | 211,391 | 250,305 | 1.18 | 38,914 | 132.1 | 32,925 | 84.6 | 97 |
| UT | 19,669 | 21,491 | 1.09 | 1,822 | 55.6 | 1,560 | 85.6 | 11 |
| VA | 71,529 | 79,893 | 1.12 | 8,364 | 97.9 | 6,252 | 74.7 | 52 |
| VT | 4,585 | 4,817 | 1.05 | 232 | 47.8 | 141 | 60.8 | 6 |
| WA | 50,831 | 53,855 | 1.06 | 3,024 | 44.4 | 3,503 | 115.8 | 18 |
| WI | 54,662 | 62,003 | 1.13 | 7,341 | 126.7 | 6,205 | 84.5 | 38 |
| WV | 22,954 | 25,247 | 1.10 | 2,293 | 132.7 | 1,601 | 69.8 | 21 |
| WY | 5,091 | 5,983 | 1.18 | 892 | 155.1 | 526 | 58.9 | 7 |

*Notes:* Aggregated results by various geographic regions. Aggregate rates are computed by summing actual and predicted counts over counties in a particular region and dividing by the summed population. Note that this is equivalent to the population-weighted means of the county-level rates. COVID-19 deaths refer to deaths that appeared as either an underlying or contributing cause on the death certificate. In line with CDC guidance,[18] we exclude counties with negative excess death rates from the aggregations. 48 county-sets representing less than 2% of the U.S. population (5.5 million residents) were excluded.

**Table A2:** U.S. Population Living in Counties with Different Proportions of Excess Deaths Assigned to COVID-19

| Percent of excess deaths assigned to COVID-19 | Number of county sets | Population | Cumulative population | Cumulative percent |
| --- | --- | --- | --- | --- |
| (5,10] | 1 | 55,379 | 55,379 | 0.02% |
| (10,15] | 1 | 58,214 | 113,593 | 0.03% |
| (15,20] | 1 | 57,834 | 171,427 | 0.05% |
| (20,25] | 5 | 341,482 | 512,909 | 0.15% |
| (25,30] | 4 | 247,713 | 760,622 | 0.23% |
| (30,35] | 5 | 422,780 | 1,183,402 | 0.36% |
| (35,40] | 6 | 528,800 | 1,712,202 | 0.52% |
| (40,45] | 12 | 1,735,533 | 3,447,735 | 1.04% |
| (45,50] | 22 | 3,224,932 | 6,672,667 | 2.02% |
| (50,55] | 45 | 5,546,759 | 12,219,426 | 3.69% |
| (55,60] | 54 | 8,418,387 | 20,637,813 | 6.23% |
| (60,65] | 77 | 14,181,616 | 34,819,429 | 10.52% |
| (65,70] | 84 | 19,064,559 | 53,883,988 | 16.27% |
| (70,75] | 93 | 37,334,323 | 91,218,311 | 27.55% |
| (75,80] | 114 | 43,347,704 | 134,566,015 | 40.64% |
| (80,85] | 98 | 31,868,535 | 166,434,550 | 50.26% |
| (85,90] | 88 | 21,619,839 | 188,054,389 | 56.79% |
| (90,95] | 84 | 23,402,967 | 211,457,356 | 63.86% |
| (95,100] | 628 | 114,164,341 | 325,621,697 | 98.34% |
| NA | 48 | 5,504,979 | 331,126,676 | 100% |

*Notes:* This table displays the number of county-sets and the population living in those county-sets with different proportions of excess deaths assigned to COVID-19. For example, 93 counties had between 70 and 75% of excess deaths assigned to COVID-19, representing 37.3 million U.S. residents. Cumulatively, 91.2 million U.S. residents live in areas with less than 75% assignment of excess deaths to COVID-19.

## 7 Appendix B: Details on Model Selection and Estimation

To select our primary specification for a distribution and link function, we computed both in-sample and out-of-sample predictive accuracy for a number of candidate specifications. Specifically, we estimated for three canonical GLM specifications for continuous and count data: Gaussian-identity, Poisson, and gamma models using data from 2010-2018.^9^ We then used the estimated model parameters to make out-of-sample predictions of 2019 mortality, and computed the mean-squared prediction error and mean absolute prediction error for each. Some prior studies have used a simple average of mortality in prior years to compute excess mortality in 2020. This has the advantage of being much simpler than fitting a high-dimensional GLM, although it will fail to capture trends, and may overweight more recent years. To gauge the improvement (if any) of using a regression model rather than a prior mean to predict deaths, we compute the predictive accuracy using both a one-year lagged value as well as a five-year prior average. **Table B1** reports the mean squared and mean absolute prediction error, both in-sample and out-of-sample, associated with each model. Unsurprisingly, all of the GLMs provide a much better fit in terms of in-sample deviations than lagged value or prior mean. The GLMs also perform better in predicting mortality rates in 2019. Overall, the Gaussian and Poisson GLMs perform similarly on both metrics, with the Poisson providing a slighly better fit both in and out of sample. Given these results, and the results of using alternative windows for estimation and prediction, we chose the Poisson specification as our primary specification.^10,11^ To obtain predicted mortality in each county set in 2020, we apply our estimated parametric conditional expectation function to 2020 data.

### 7.1 Uncertainty Intervals

We also construct a 95% prediction interval around the predicted death rates for 2020 to help identify counties in which 2020 mortality falls outside the normal range of year-to-year fluctuations. Importantly, while the point estimates of predicted mortality do not depend on the particular variance function assumed, the prediction intervals depend on both the variance of estimated parameters and the variance of outcome variable.^12^ To compute the prediction intervals, we first compute cluster-robust standard errors (Wooldridge, 1999)[35] for the individual parameter estimates, and then perform a parametric bootstrap procedure over the parameter and outcome distributions.^13^

**Table B1:** Prediction Accuracy of Different Models

|  | In Sample |  | Out-of-Sample |  |
| --- | --- | --- | --- | --- |
|  | Mean Sq. Err. | Mean Abs. Err. | Mean Sq. Err. | Mean Abs. Err. |
| Prior Year | 0.282 | 0.400 | 0.274 | 0.391 |
| Prior 5-Year Mean | 0.329 | 0.444 | 0.340 | 0.455 |
| Gaussian-Identity GLM | 0.108 | 0.246 | 0.216 | 0.348 |
| Poisson GLM | 0.104 | 0.241 | 0.215 | 0.347 |
| Gamma GLM | 0.106 | 0.244 | 0.229 | 0.359 |

*Notes:* Mean squared error and mean absolute error of different statistical models of mortality. Models are estimated on annual mortality data from the CDC Wonder database once from 2009 to 2017 and once from 2010 to 2018. Predictions and out-of-sample errors are then generated for 2018 and 2019 and averaged.

**Table B2:** Prediction Accuracy of Different Models

| Window | Mean Sq. Err |
| --- | --- |
| 9 | 0.215 |
| 8 | 0.221 |
| 7 | 0.229 |
| 6 | 0.244 |
| 5 | 0.262 |
| 4 | 0.278 |

*Notes:* Mean squared error of poisson GLM applied to different lookback windows.

## 8 Appendix C: All Estimates

Table C1 below includes all of our primary estimates for each county-set. Mortality rates are in units per 100,000 person-years. All-cause and COVID-19 deaths are based on provisional data from the National Center for Health Statistics. Lower CI and upper CI denote the approximate lower and upper bounds on the 95% prediction interval around expected and excess deaths in 2020. Note that the ratio of COVID-19 to excess deaths is negative in some cases, indicating that the point estimate on excess deaths is negative for the county-set. In these cases the ratio does not have a clear interpretation.

**Table C1:**
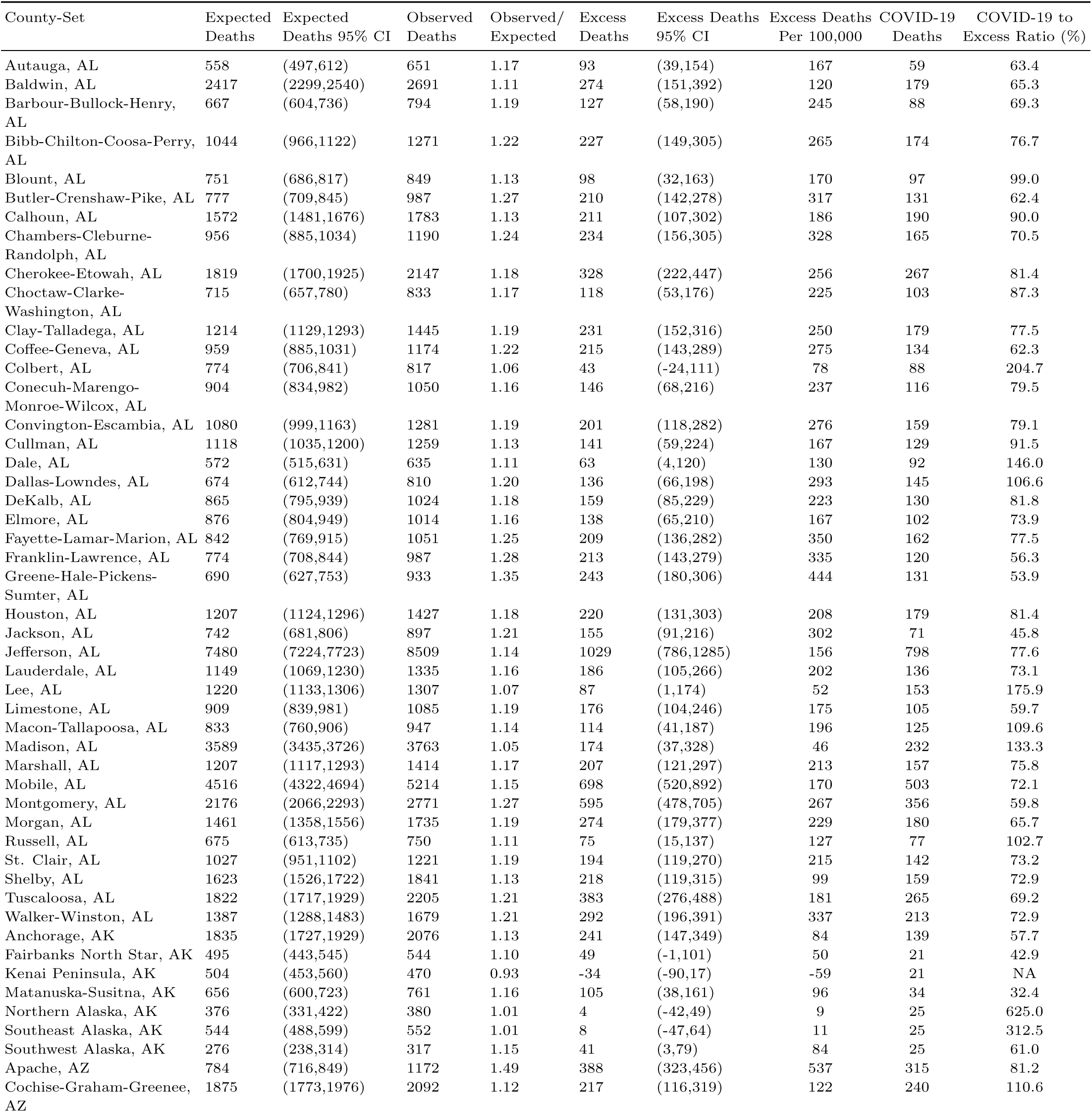
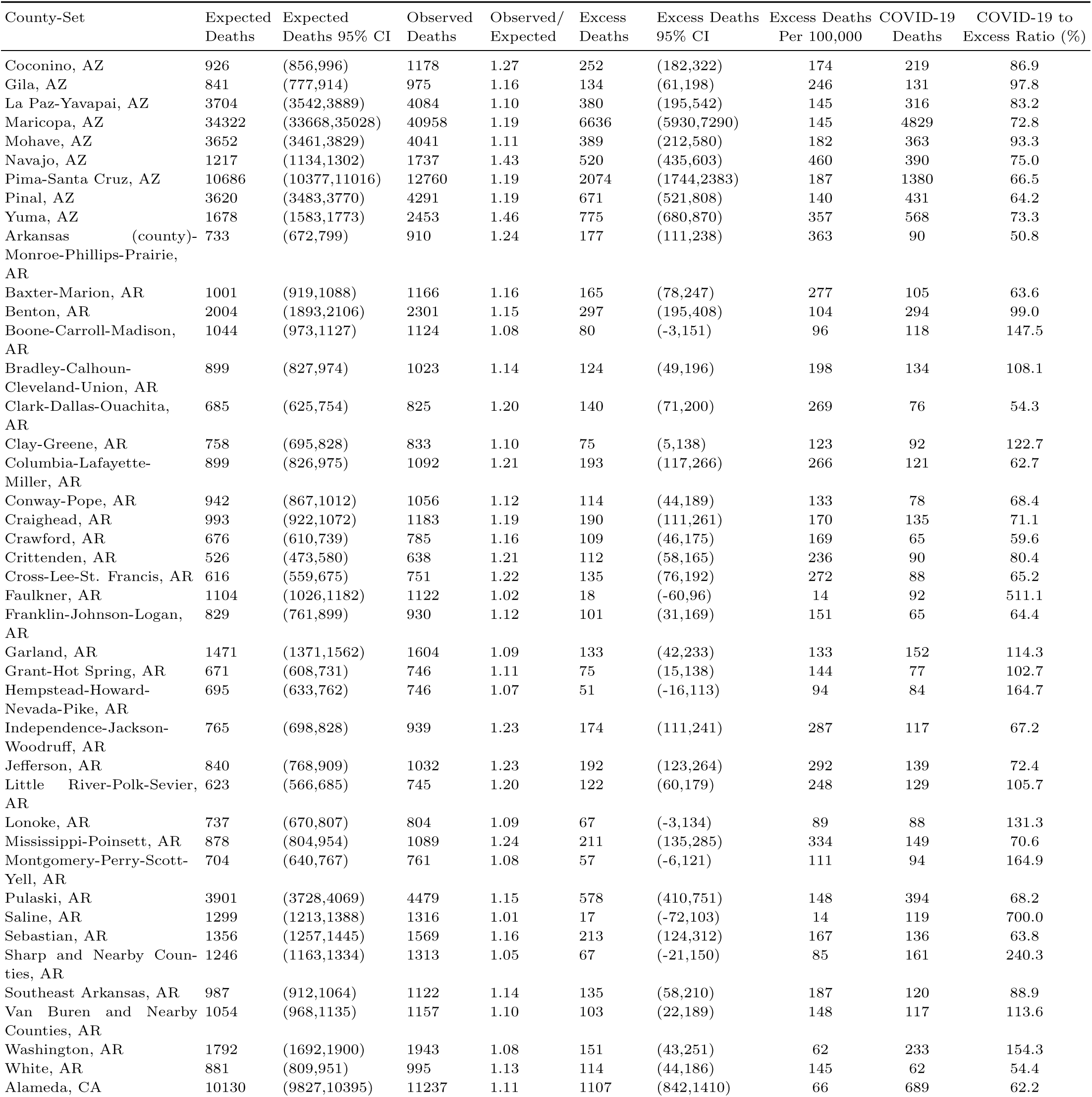
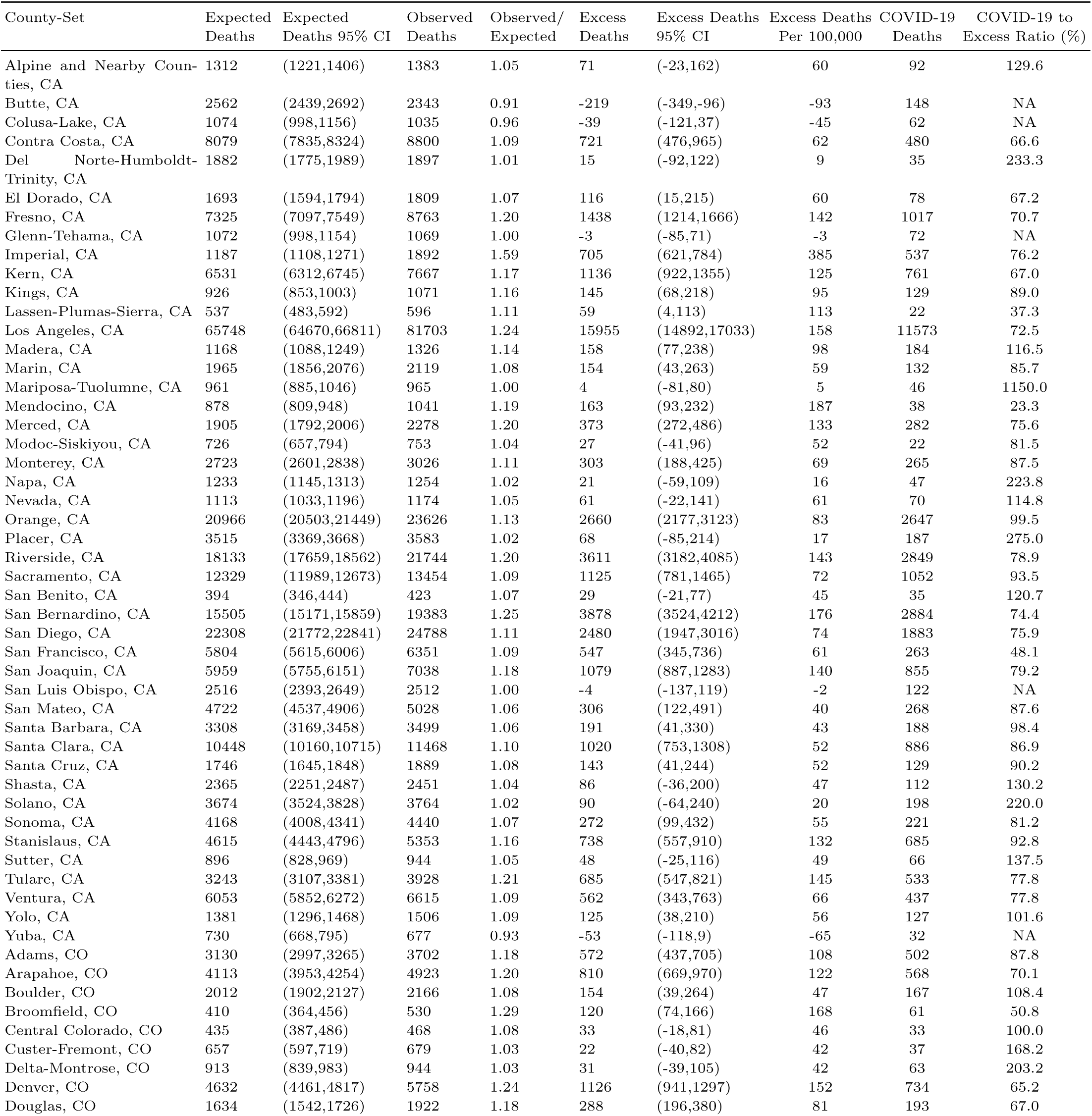
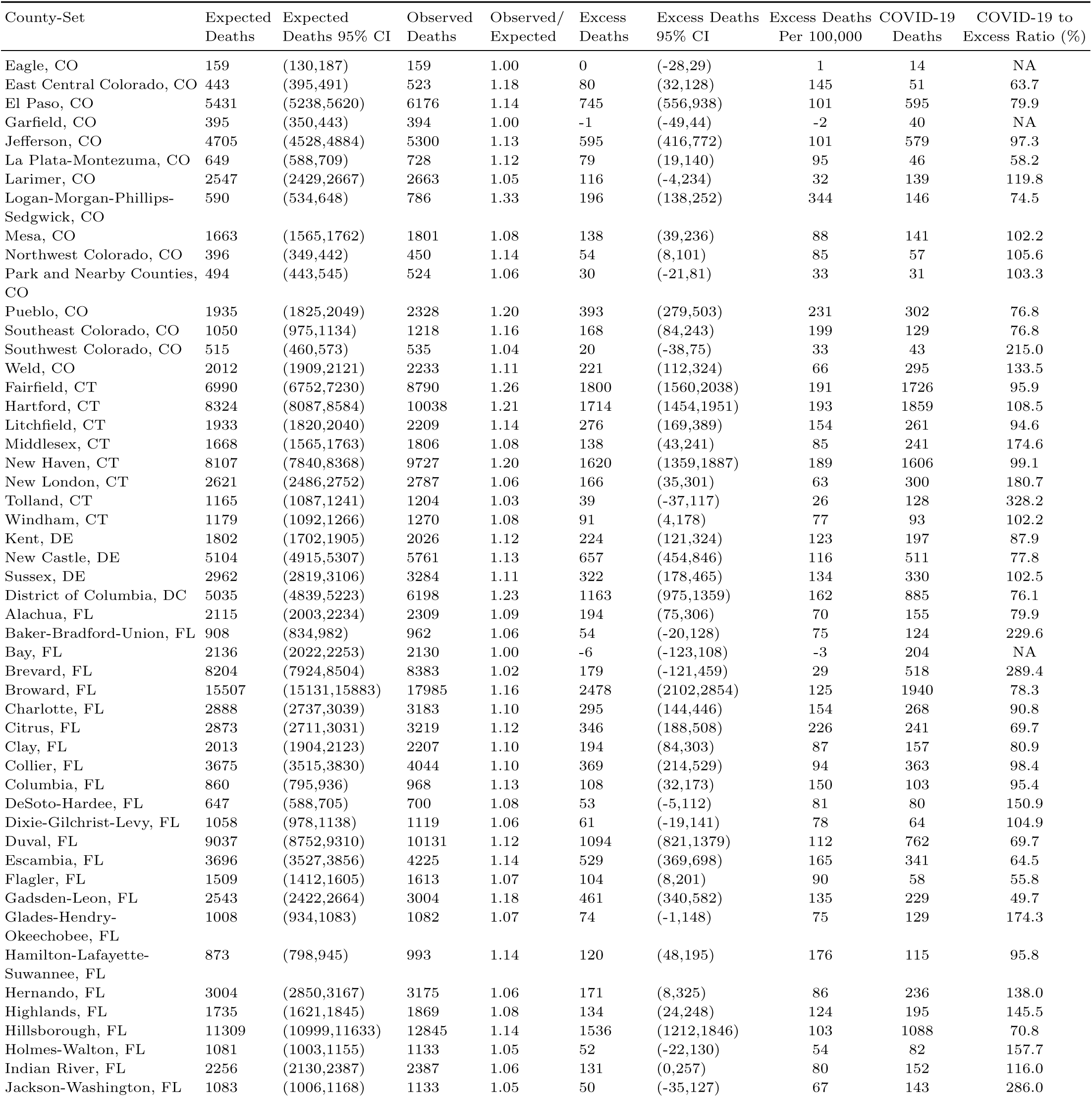
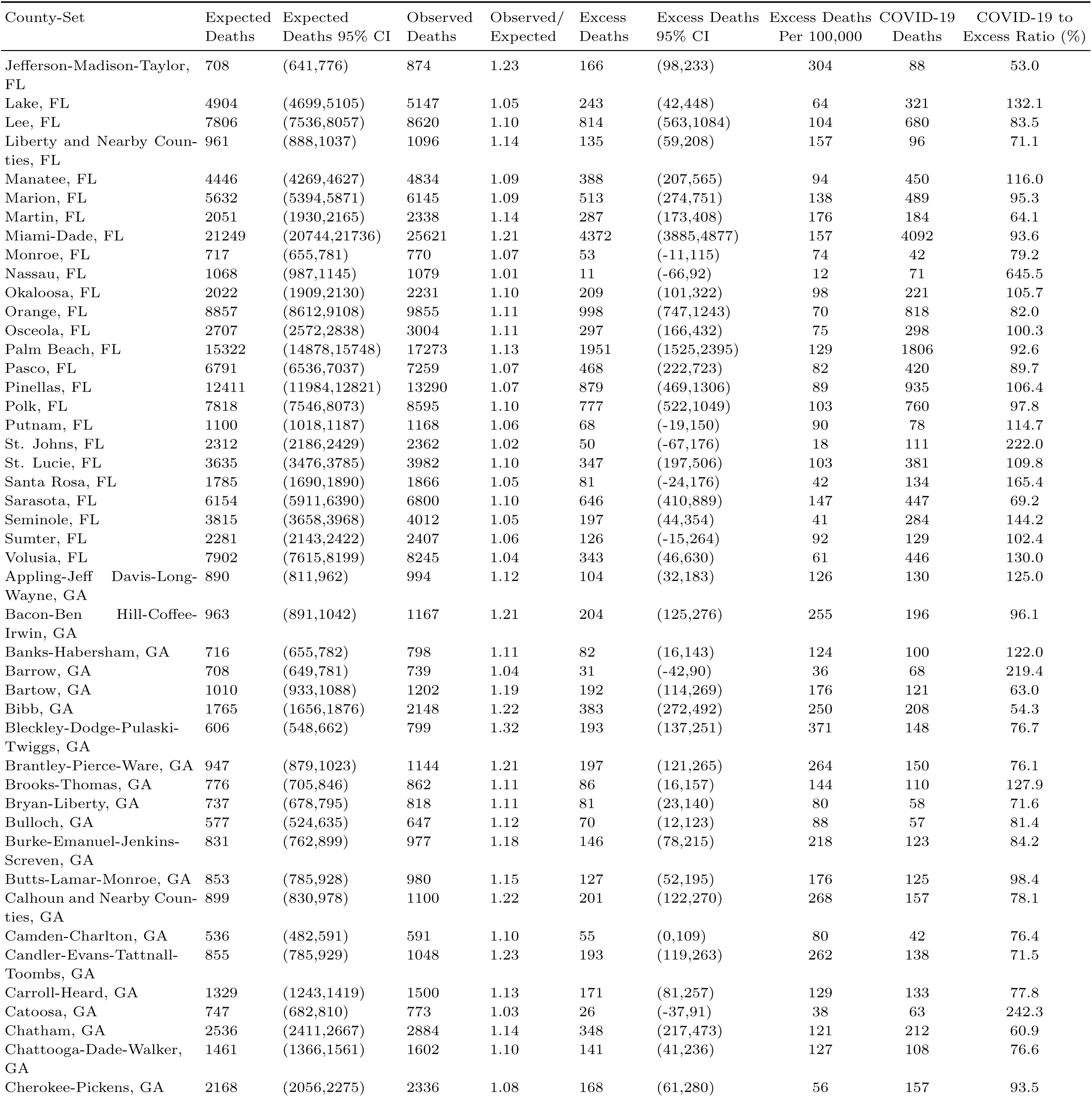
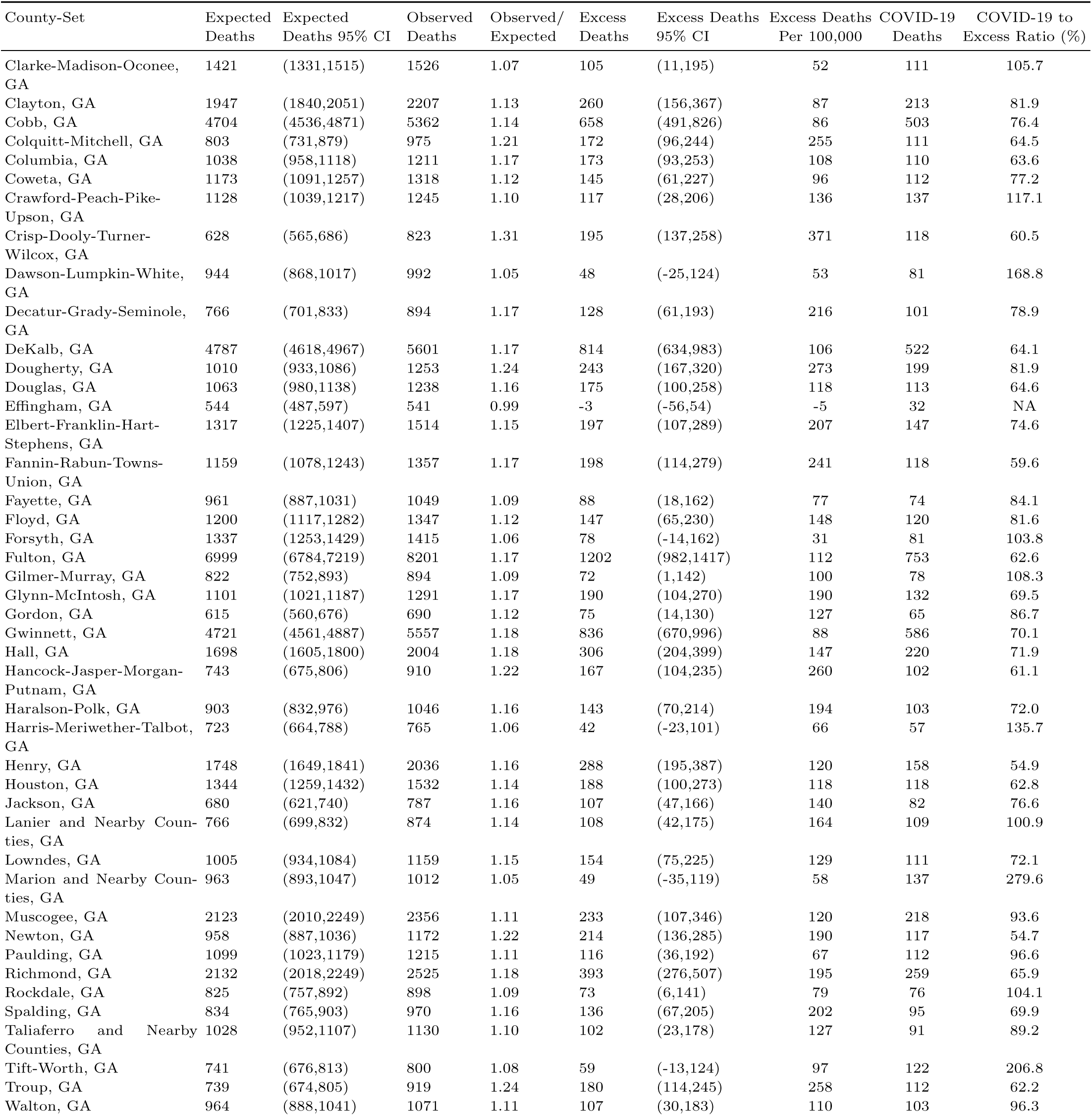
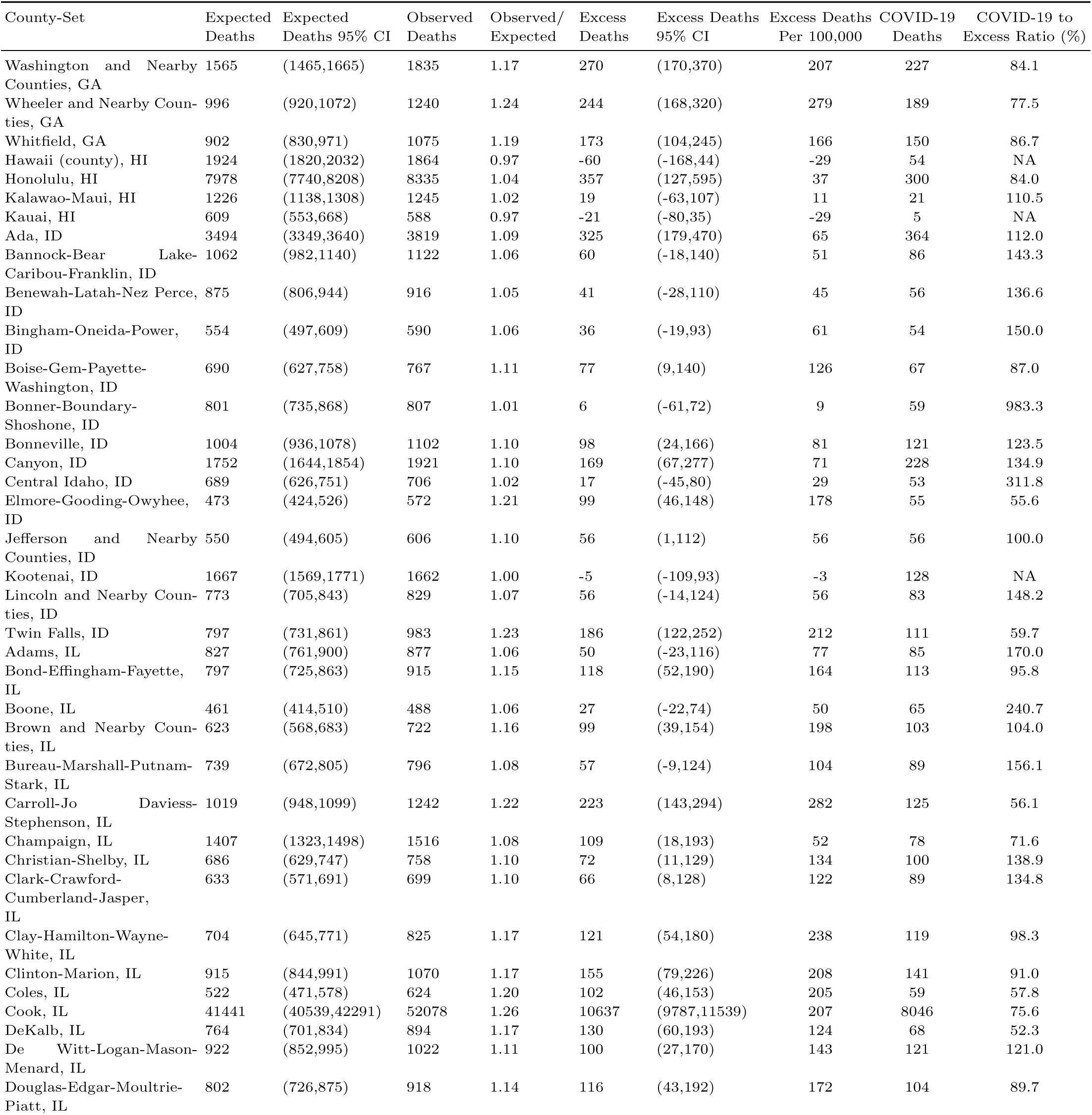
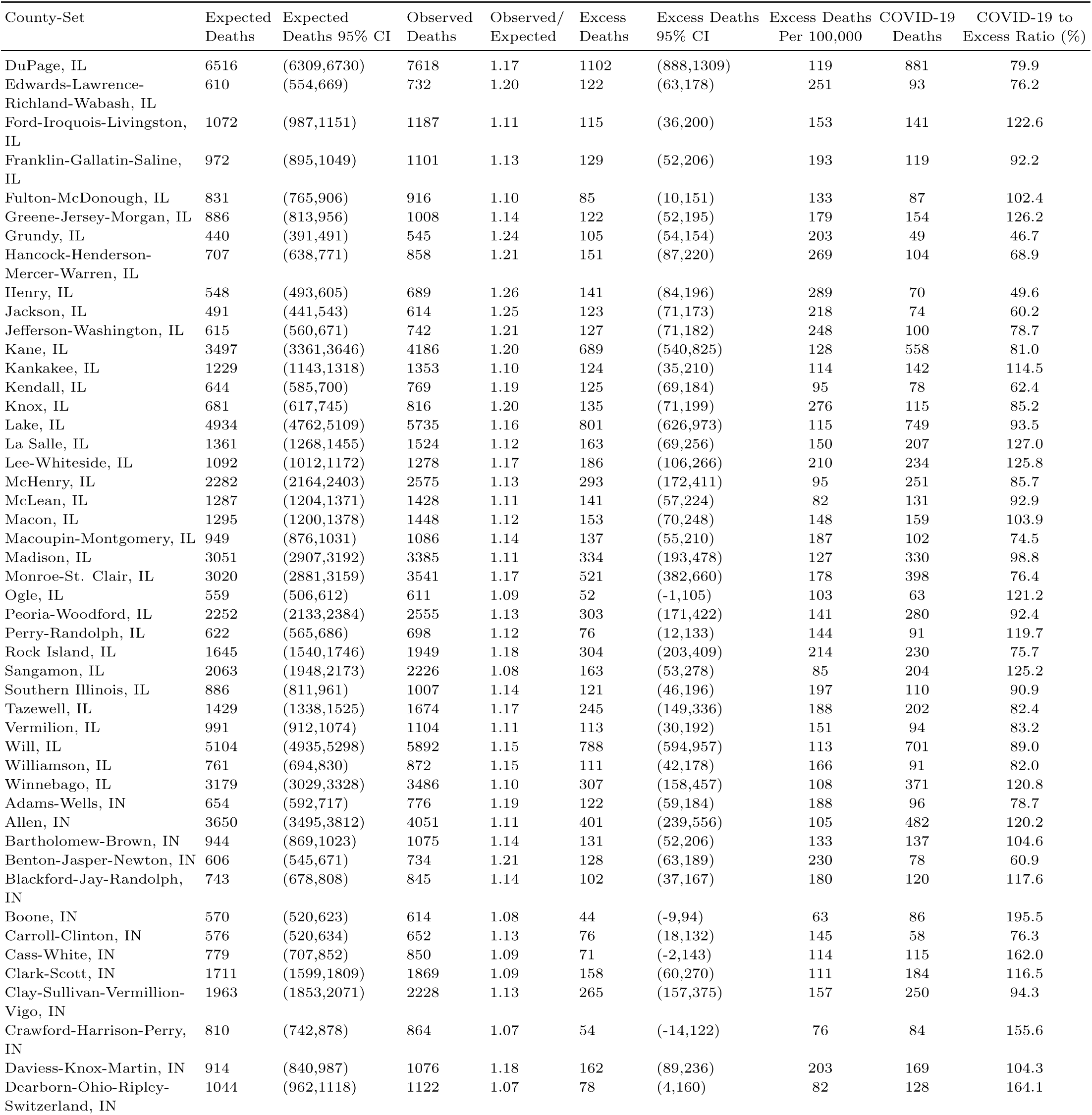
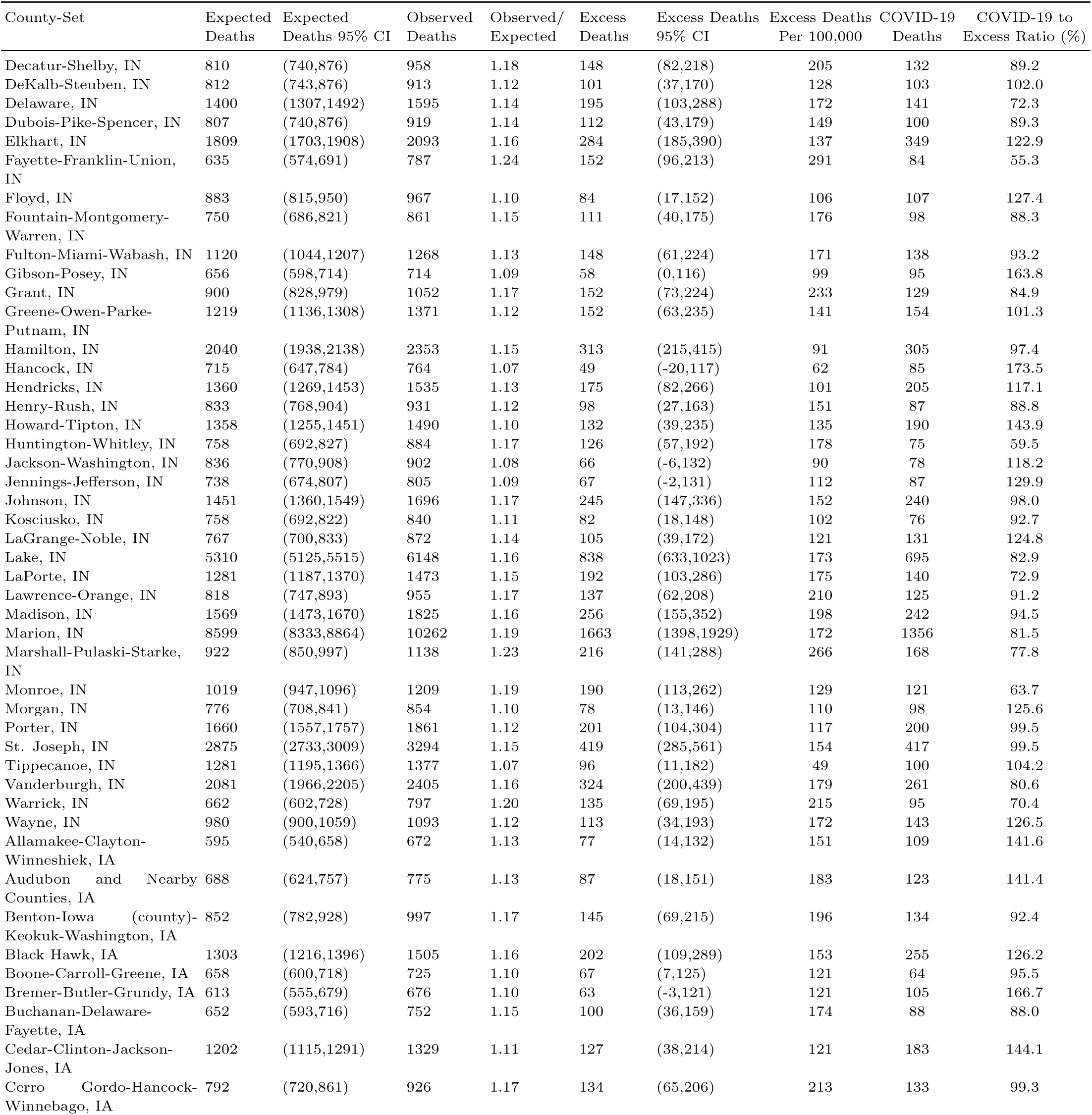
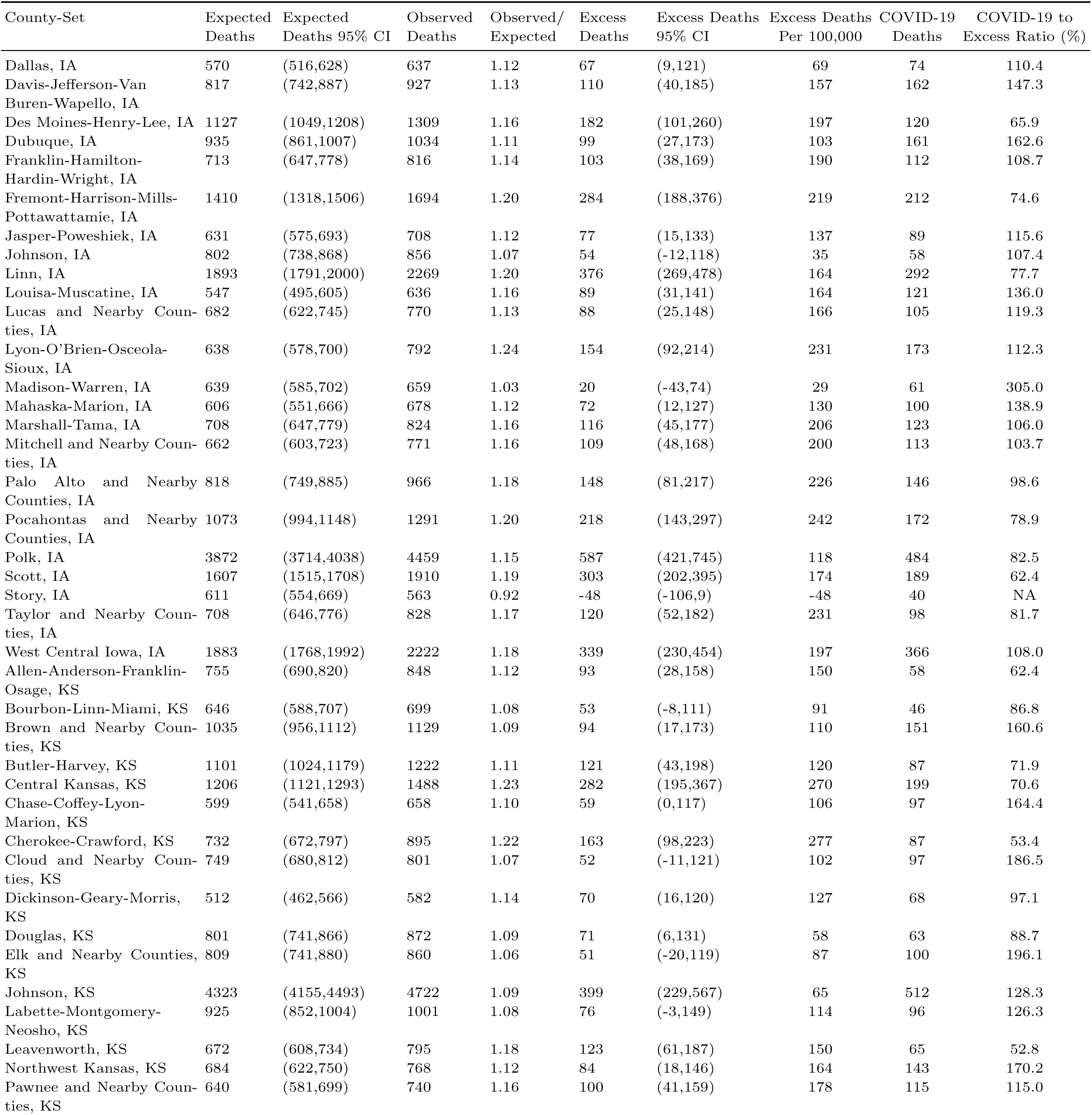
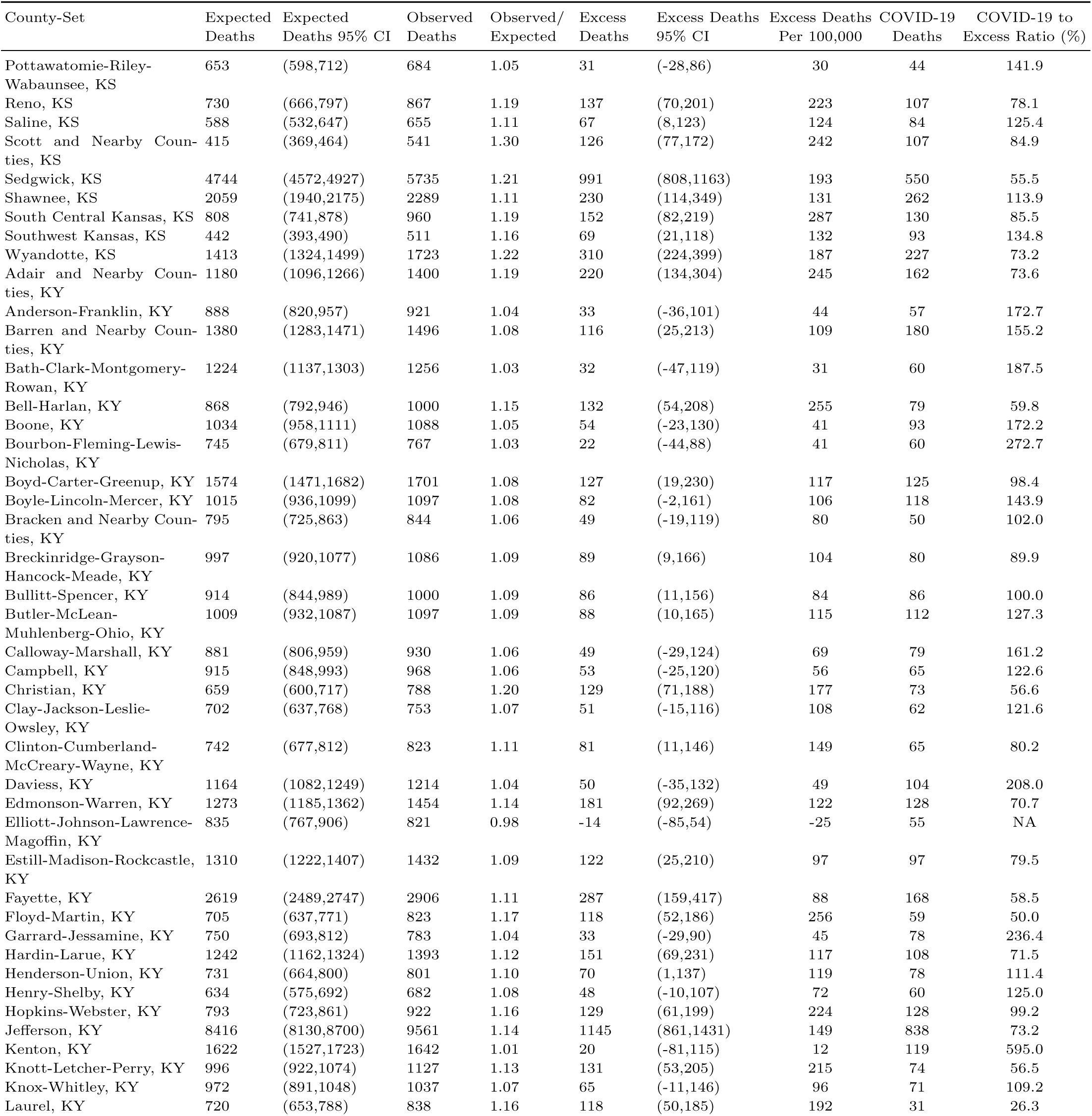
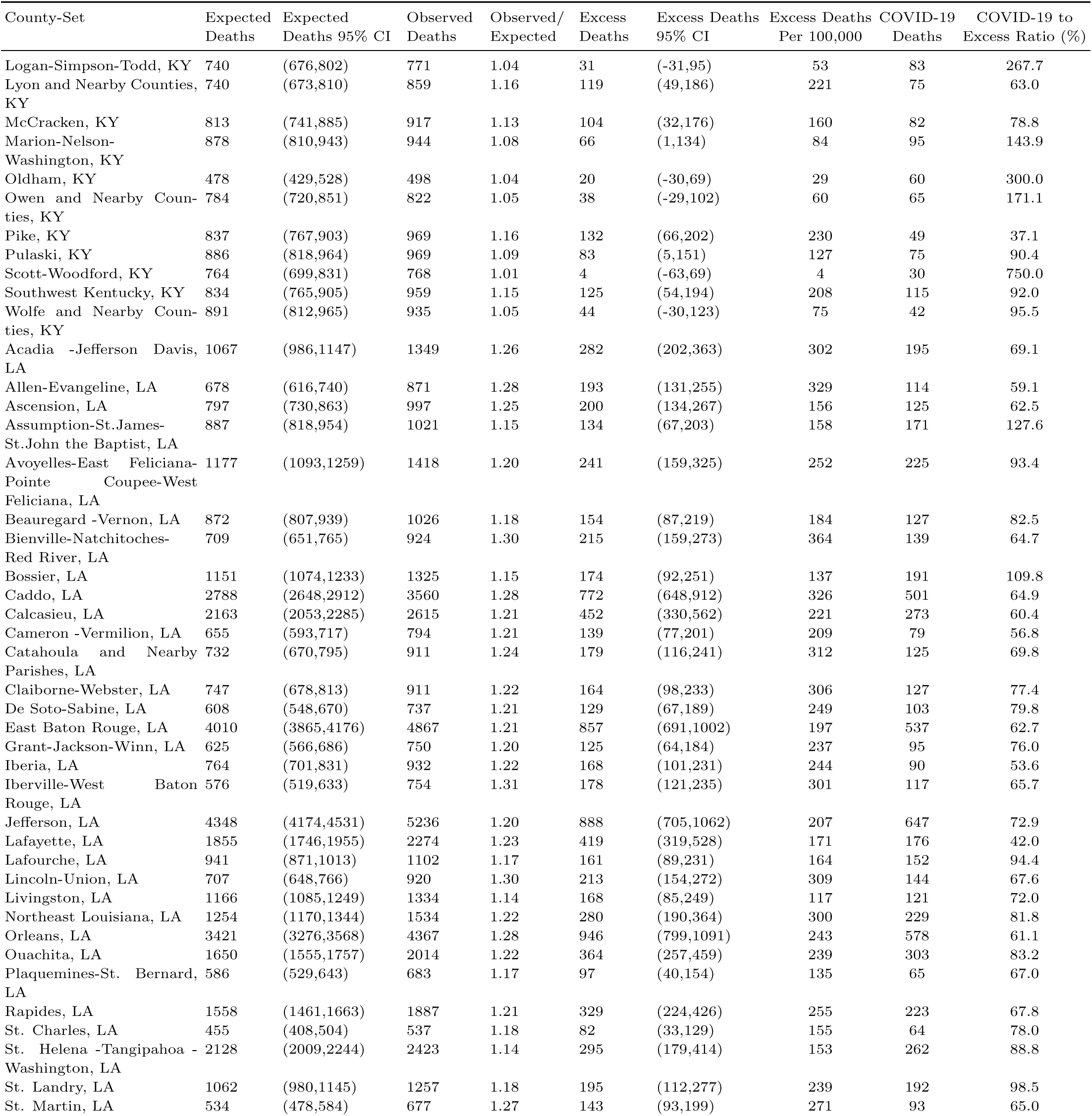
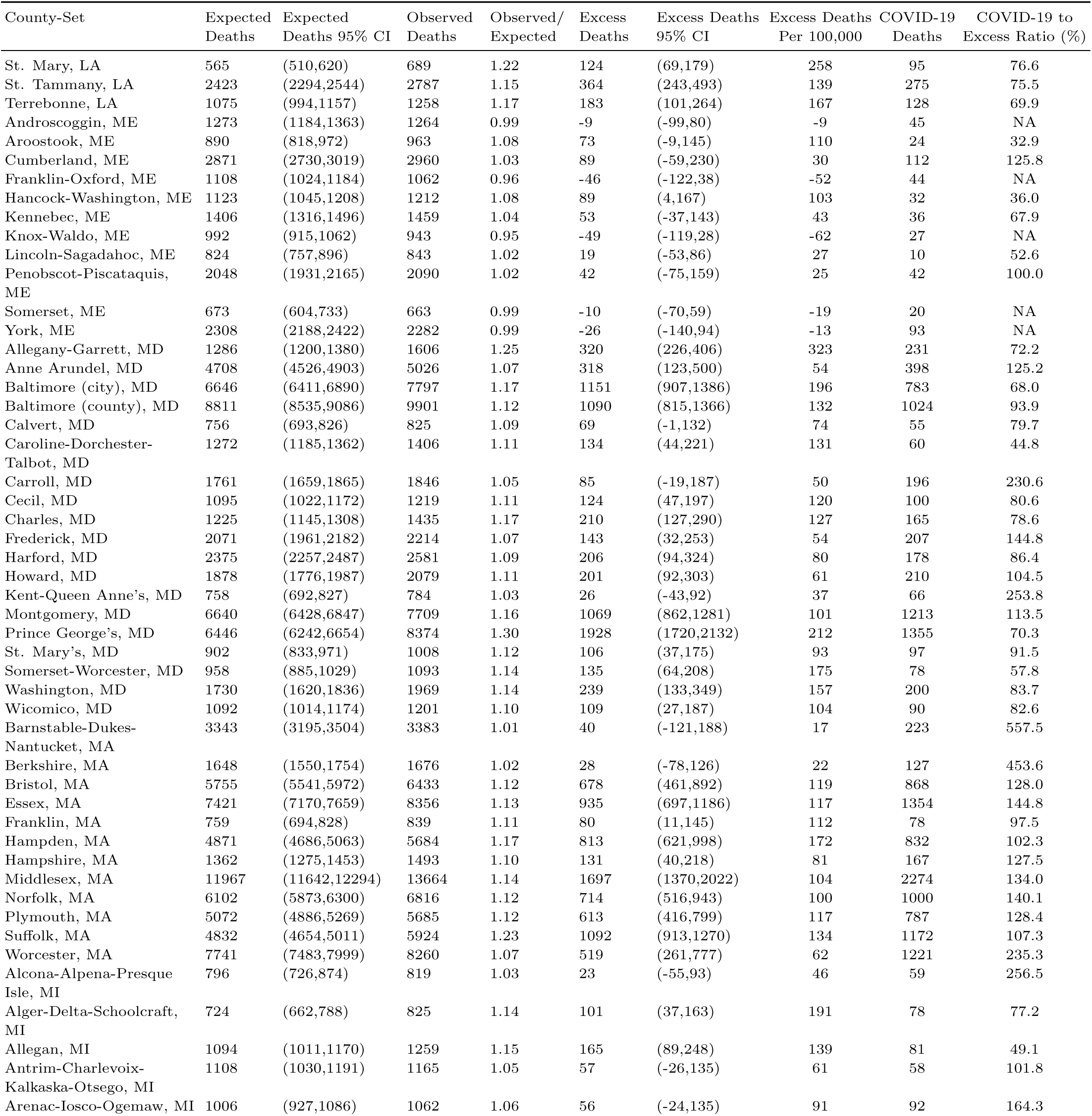
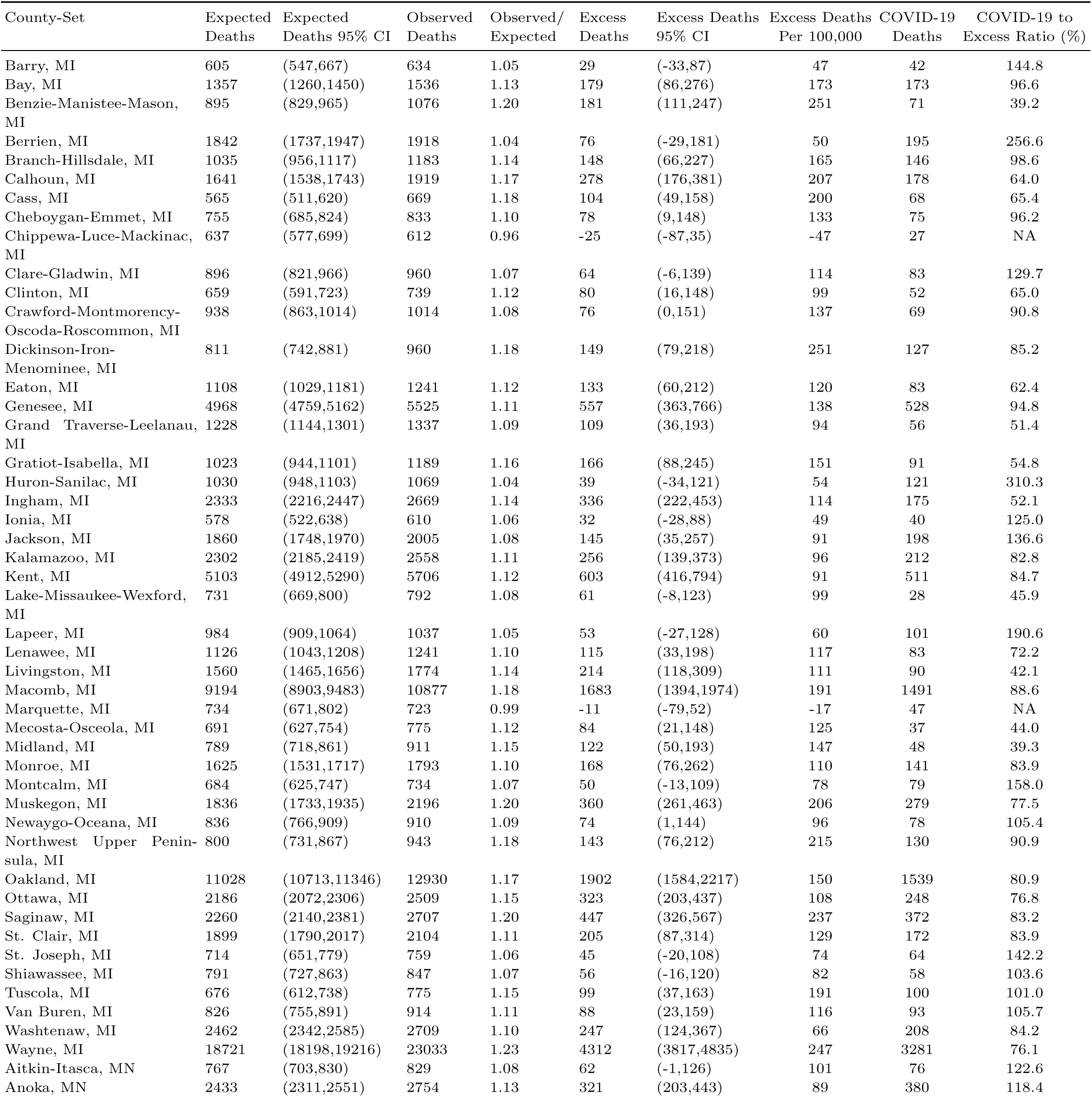
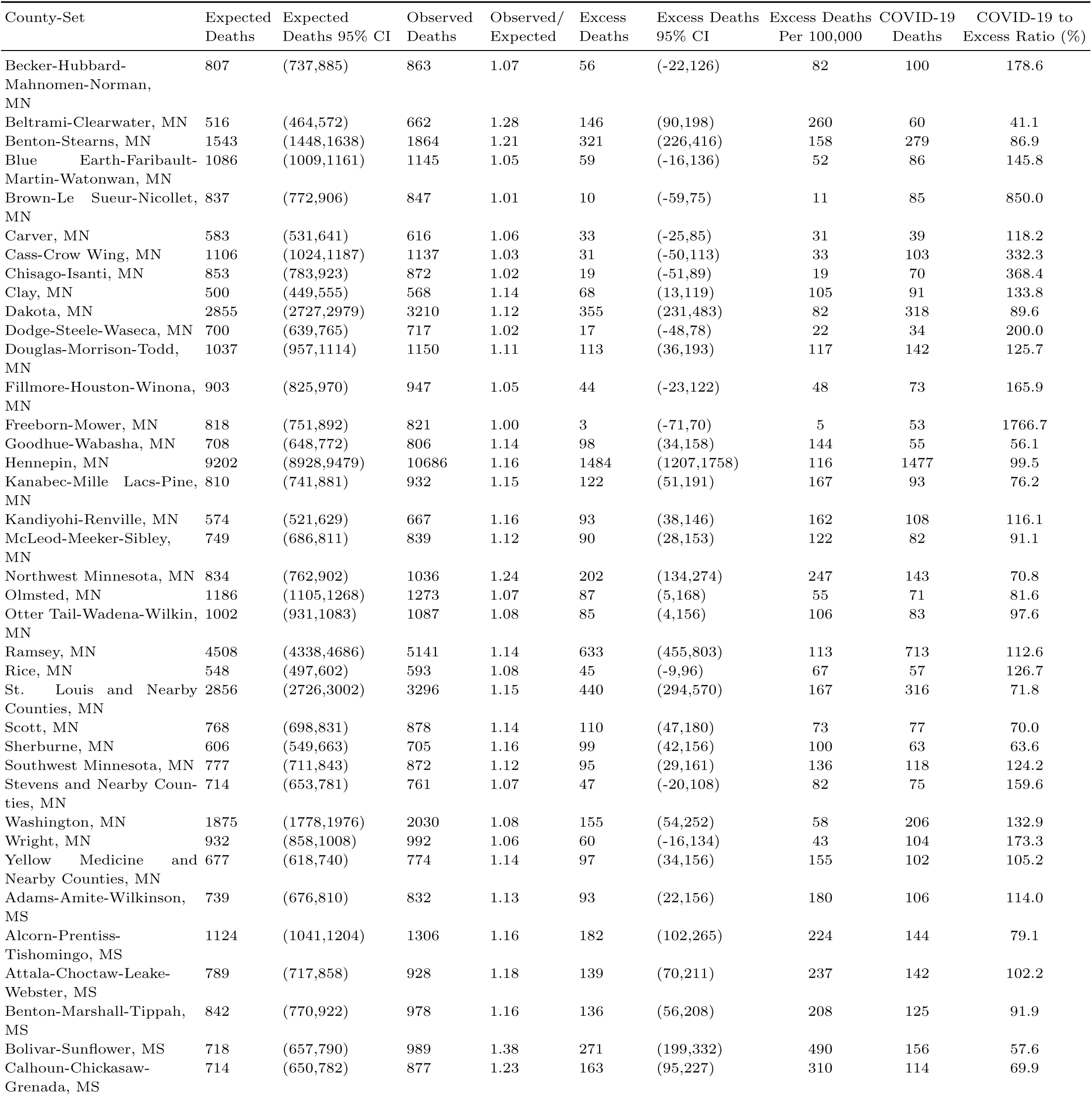
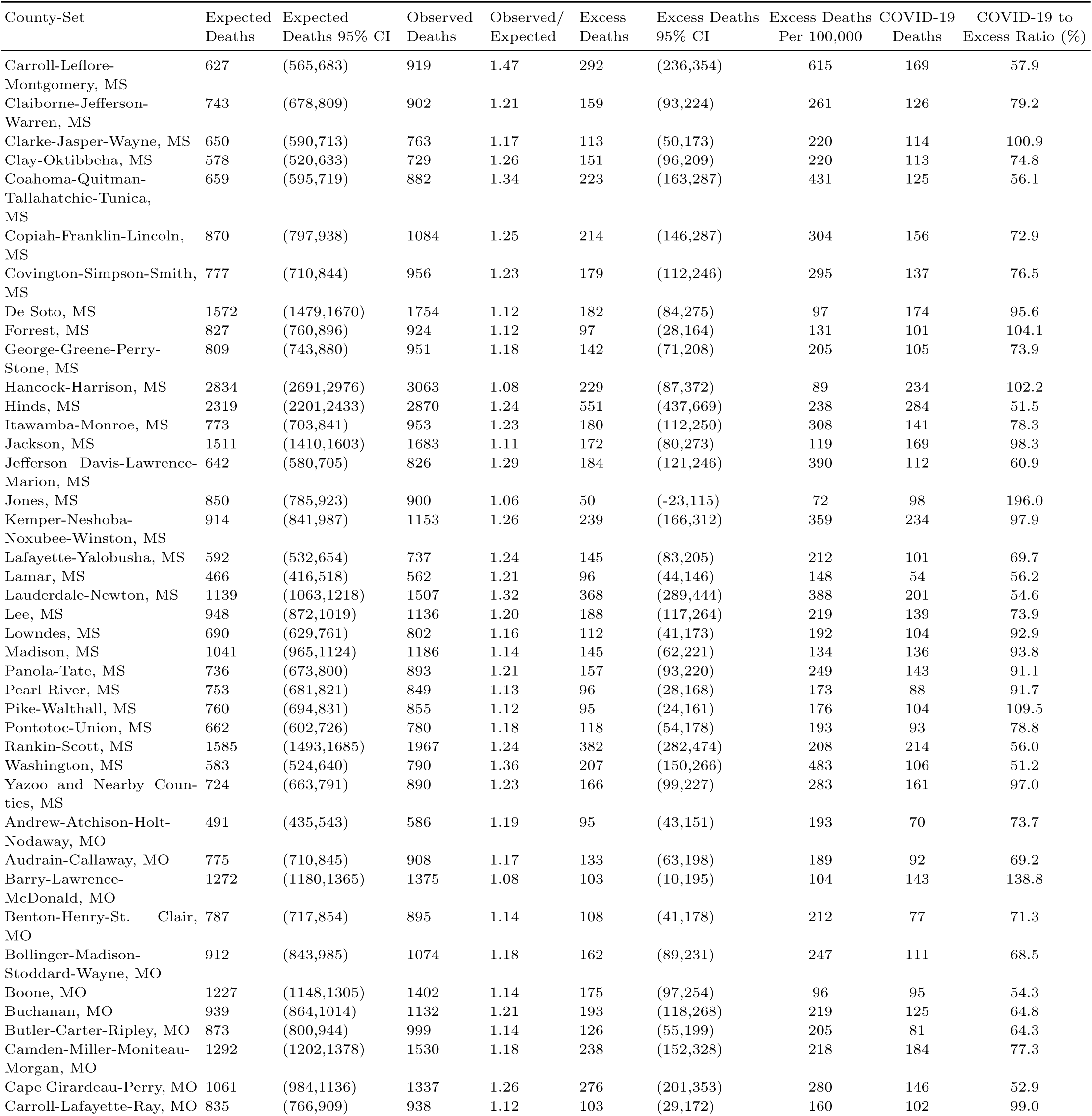
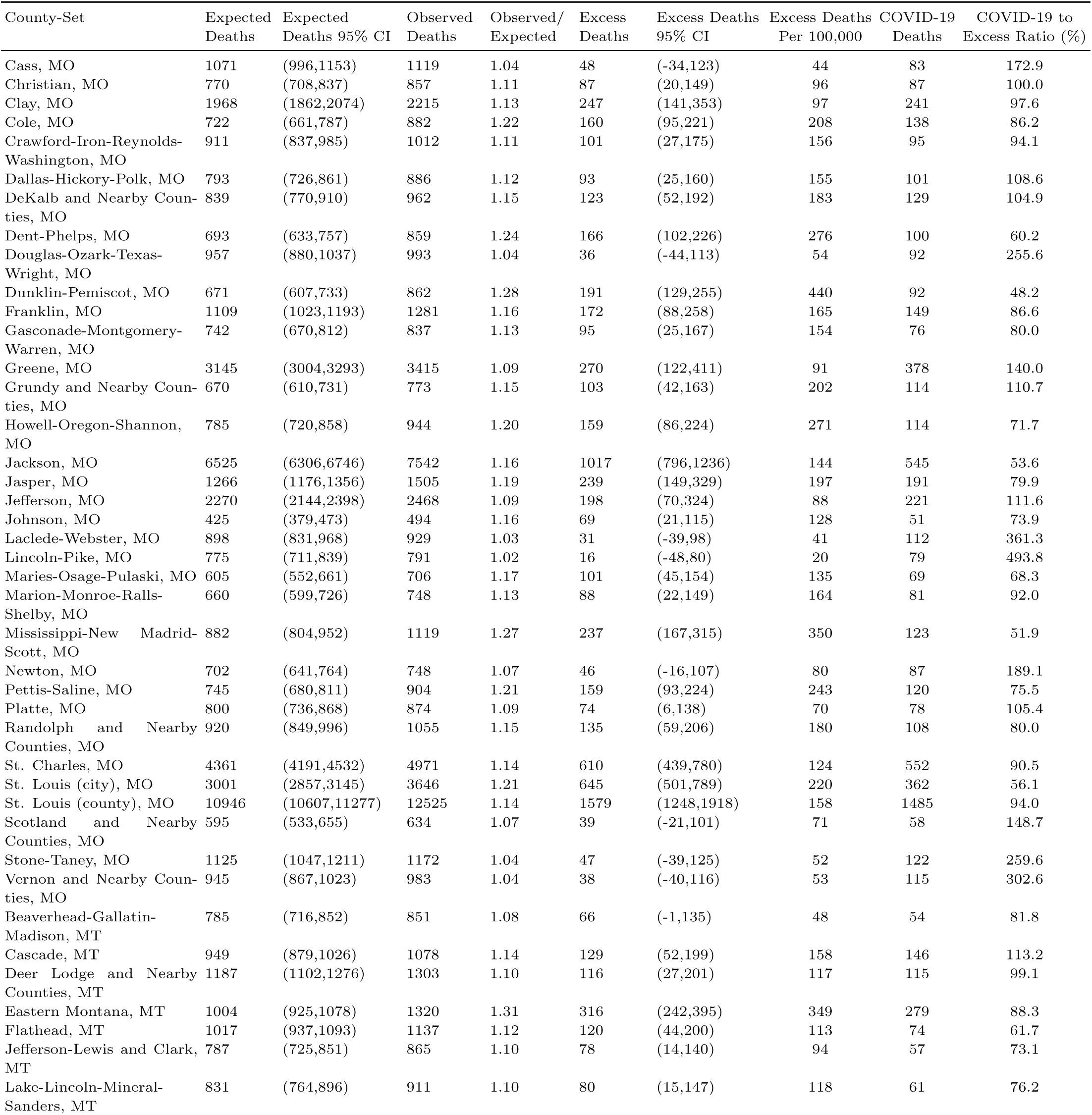
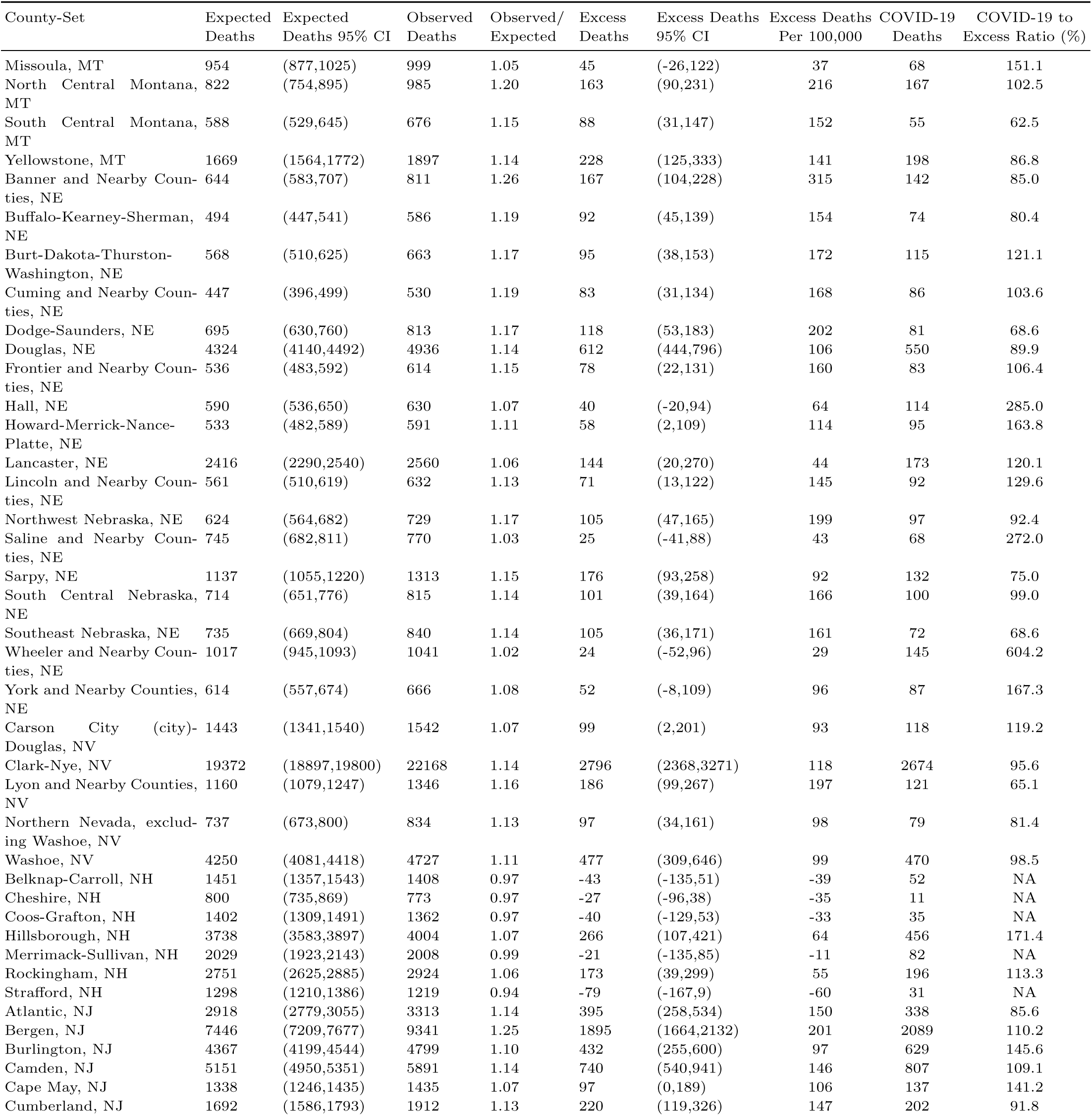
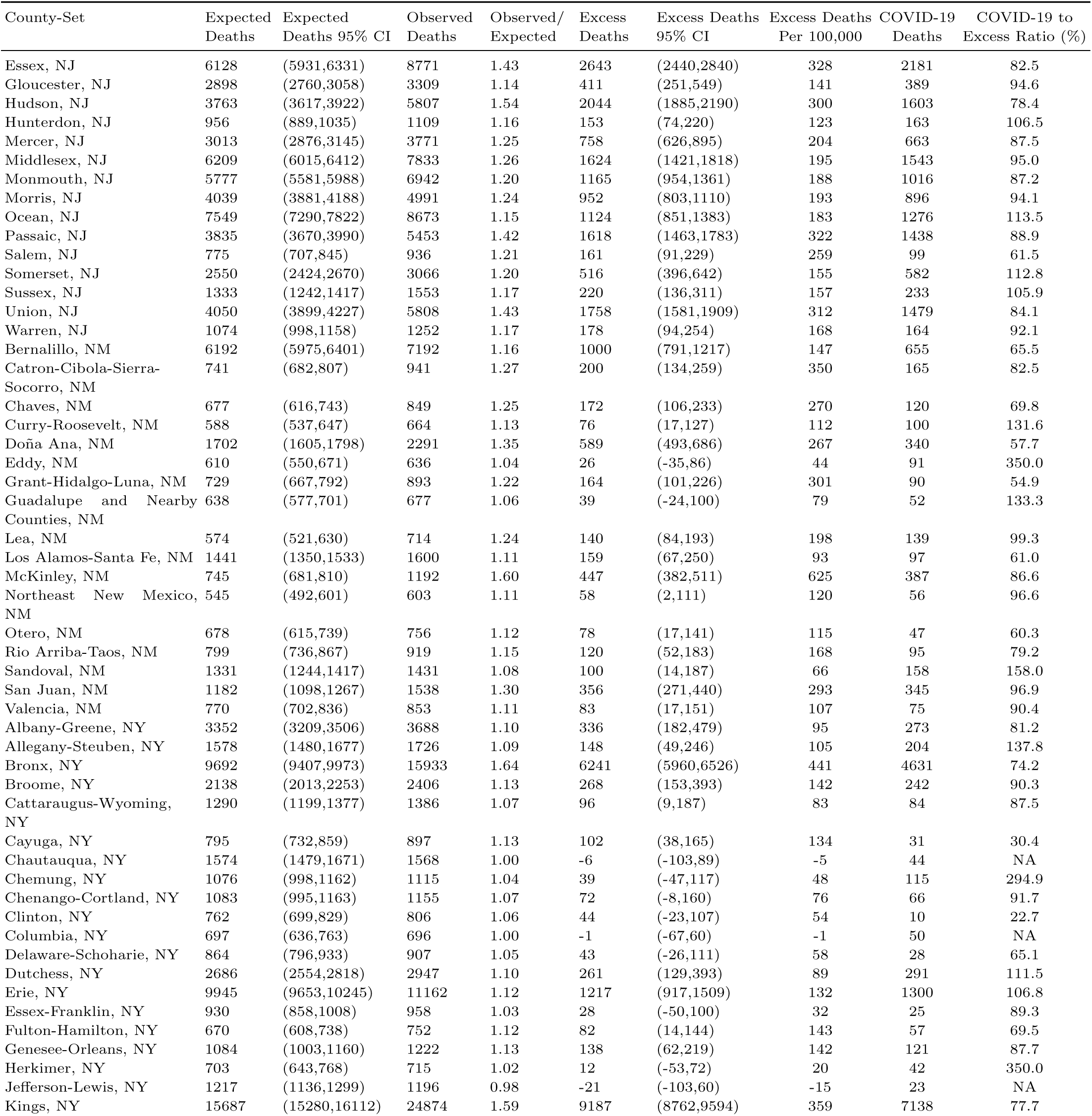
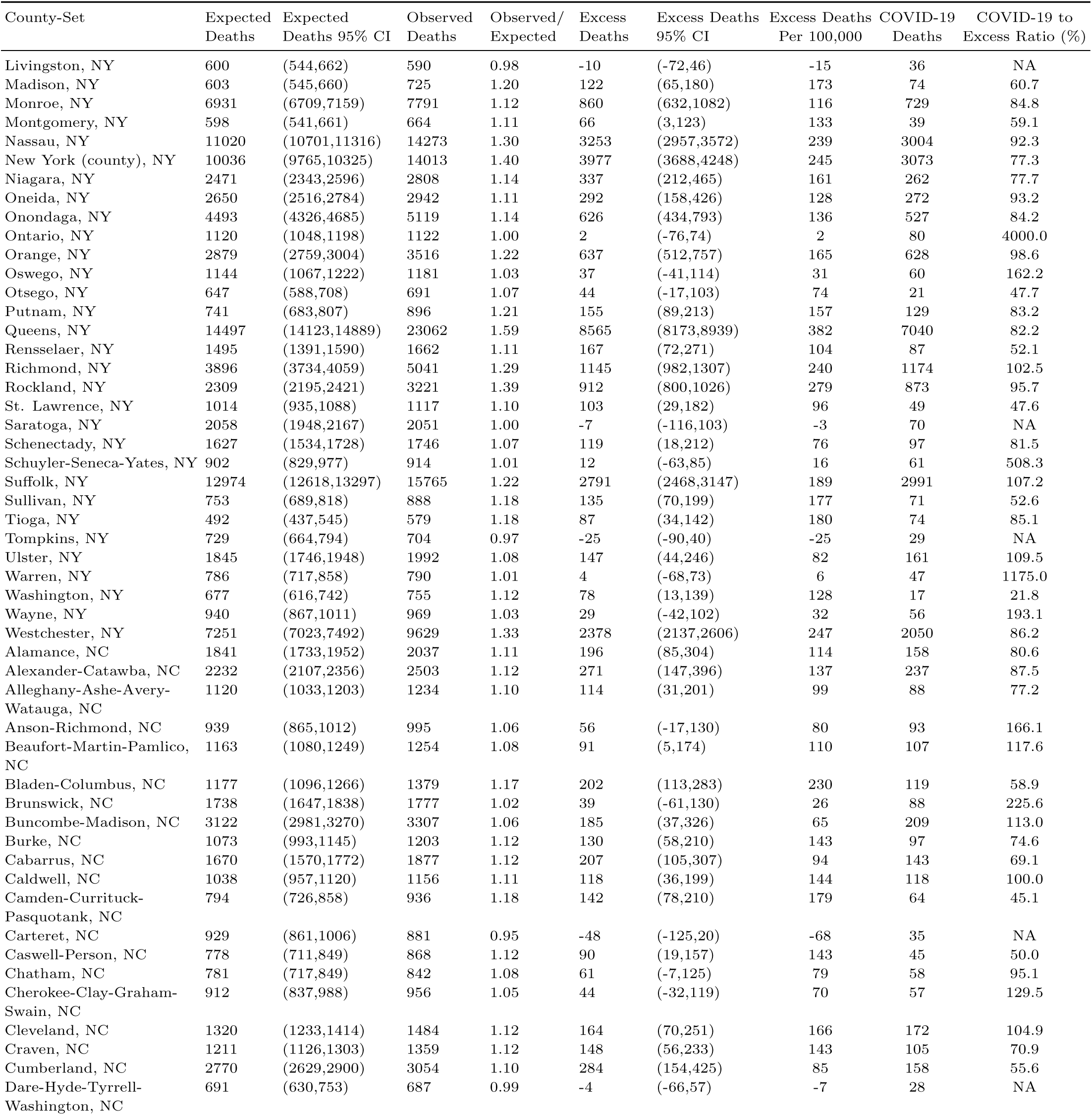
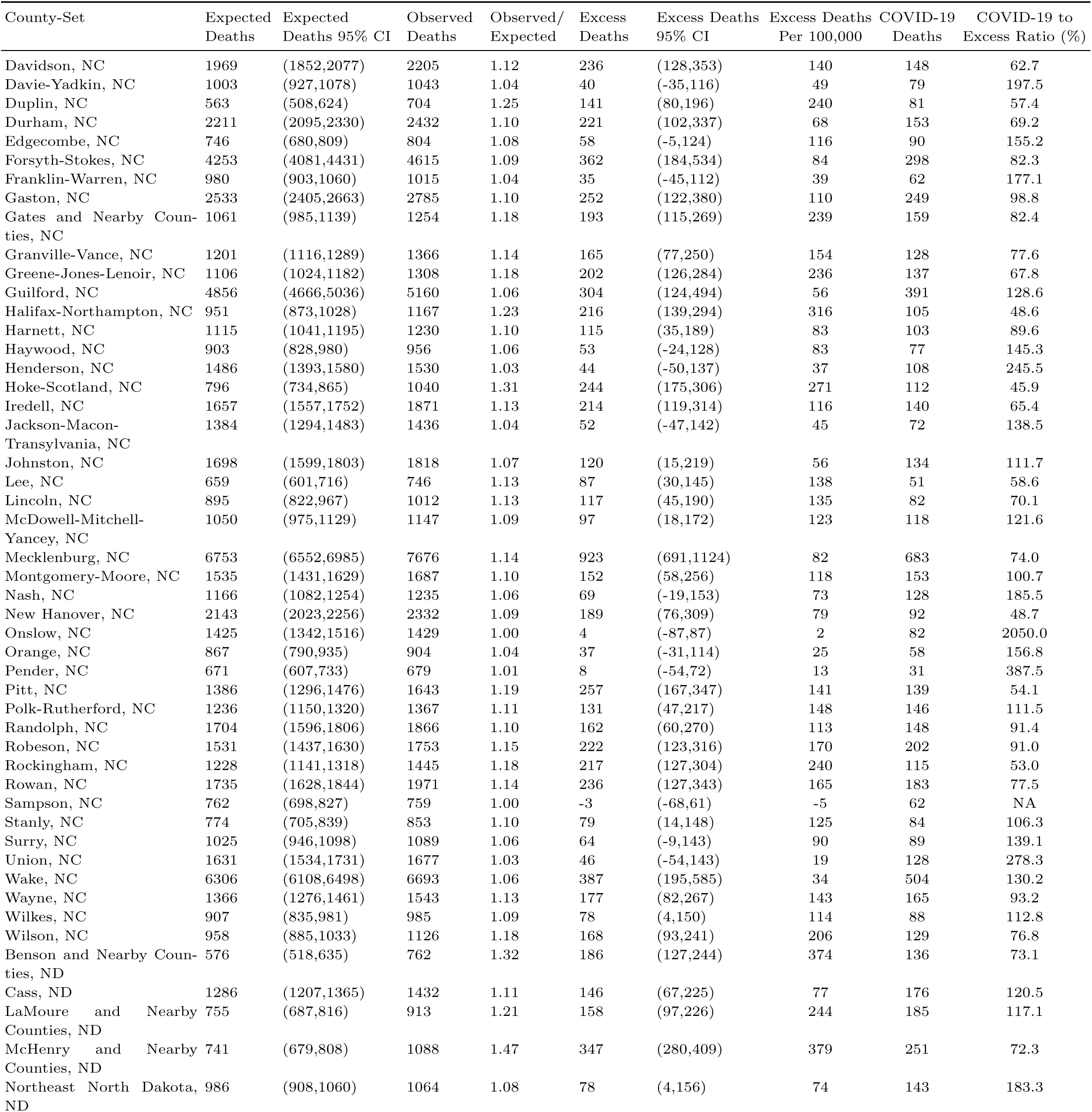
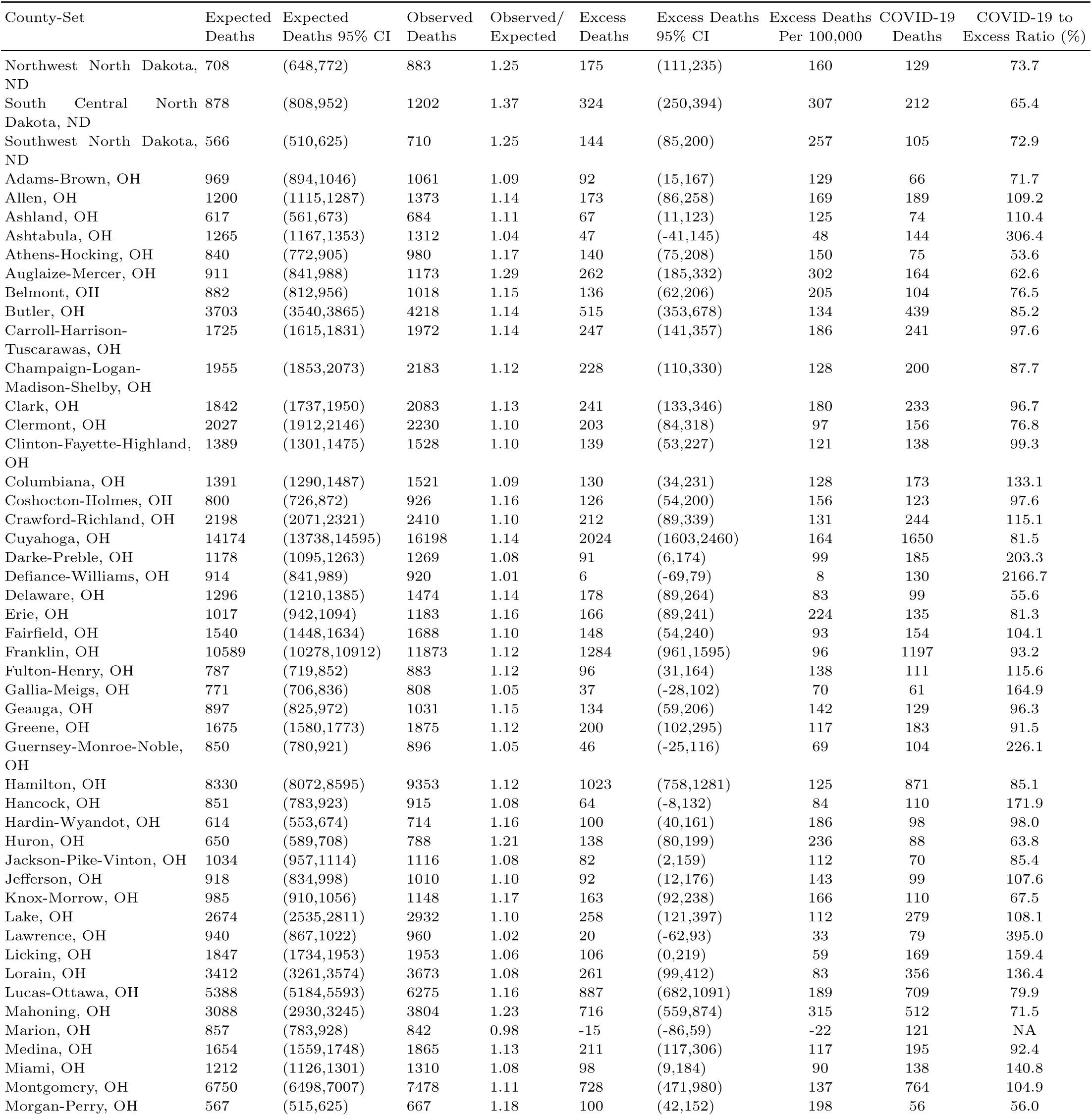
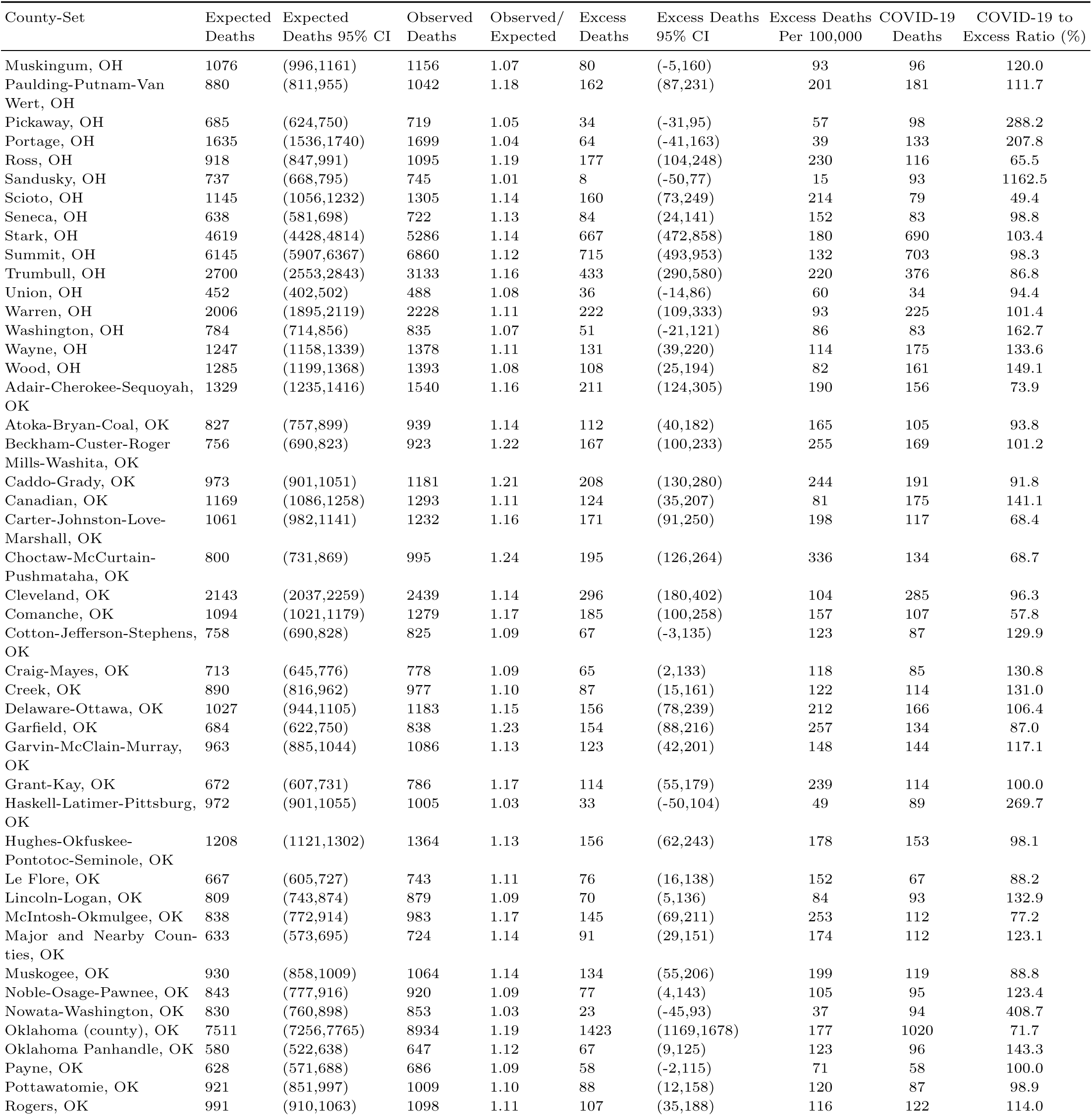
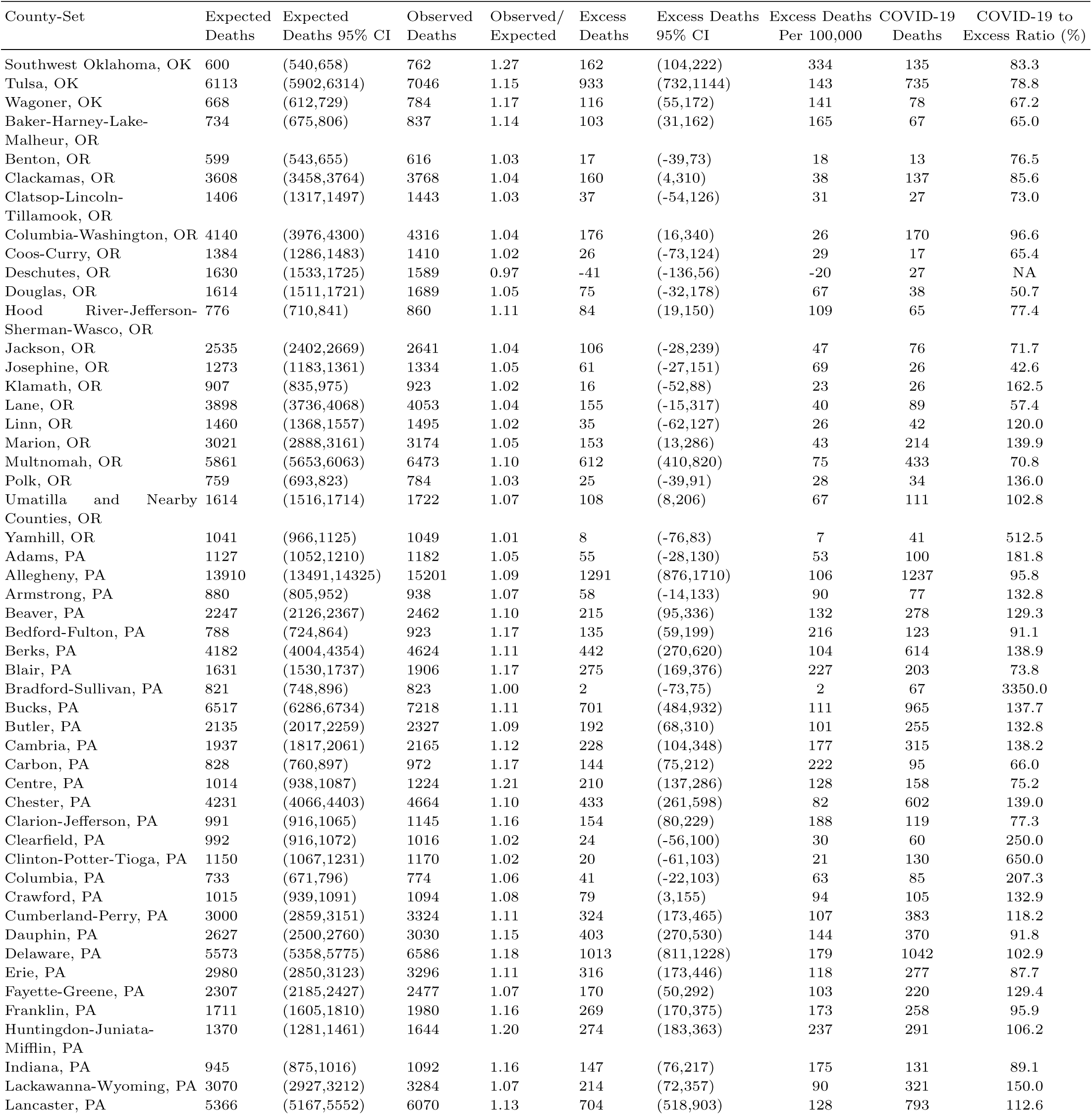
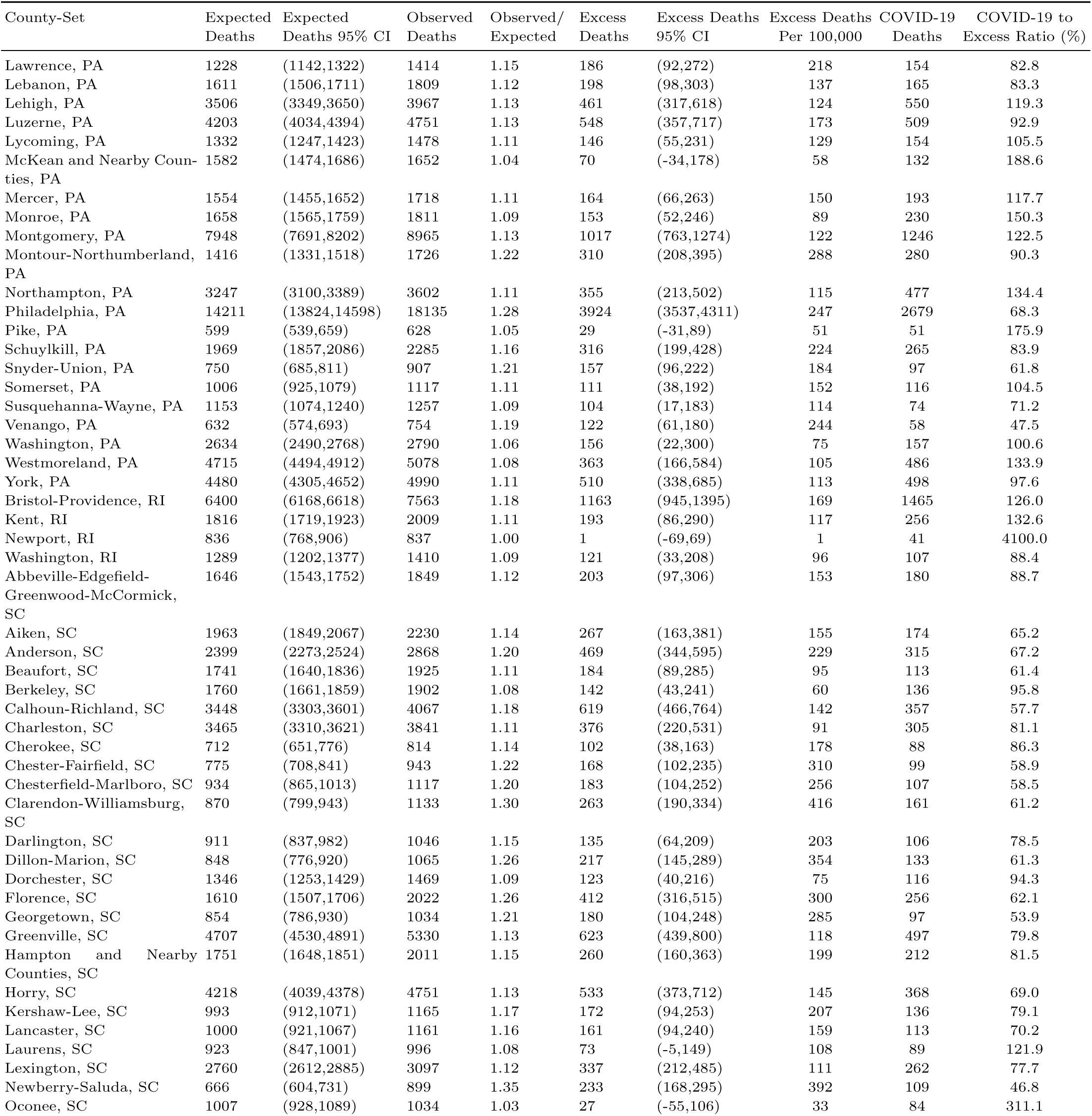
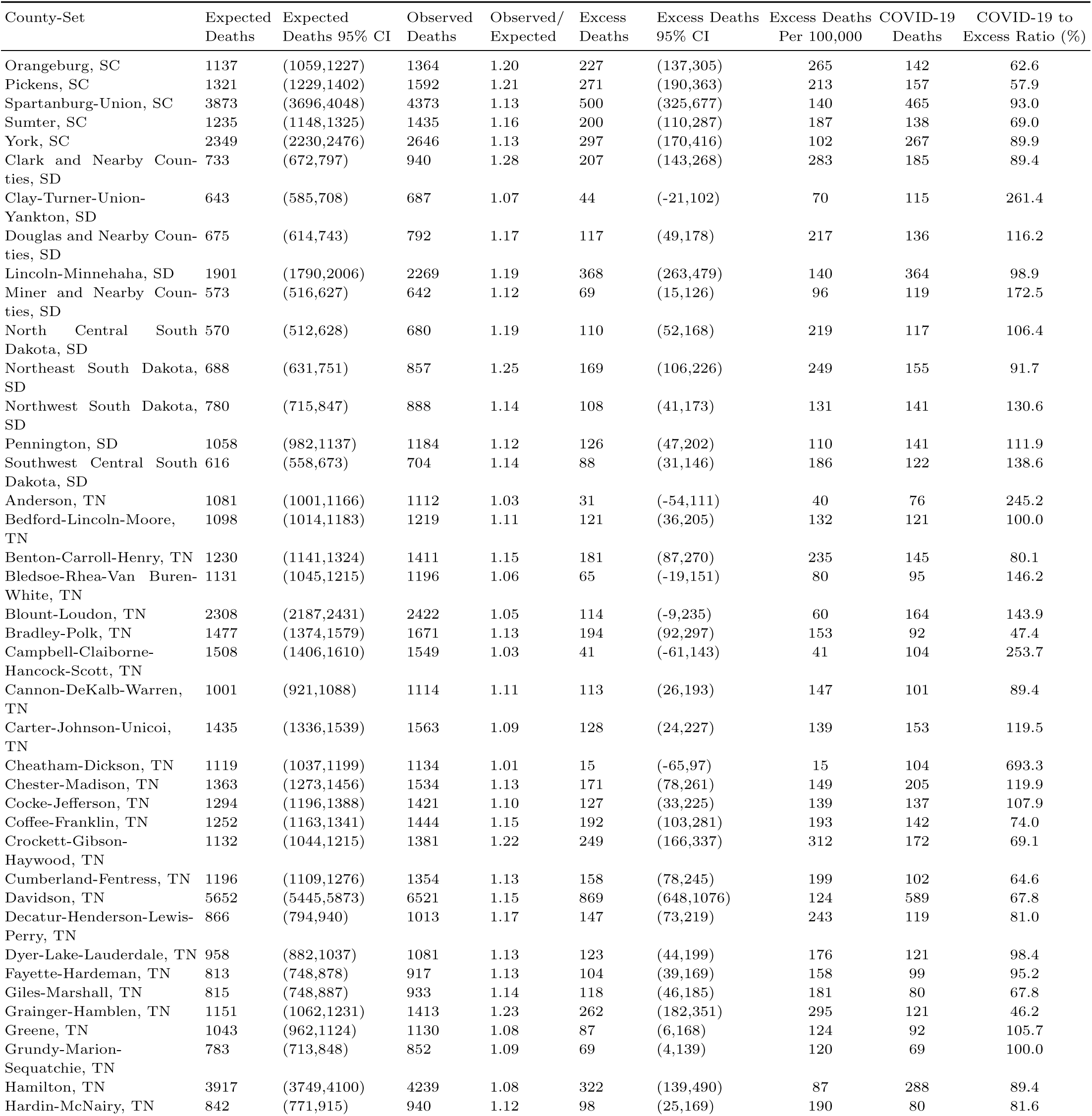
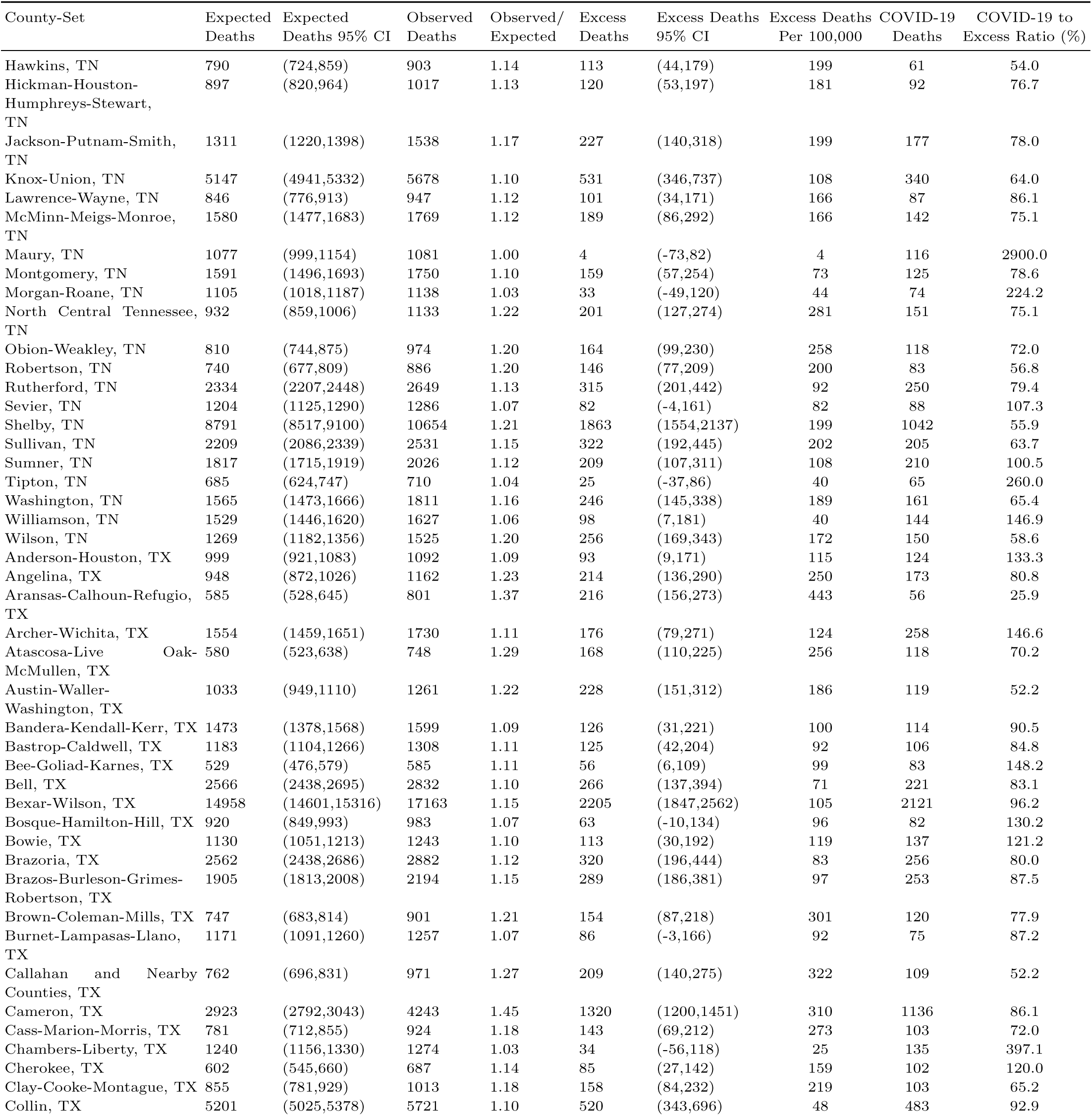
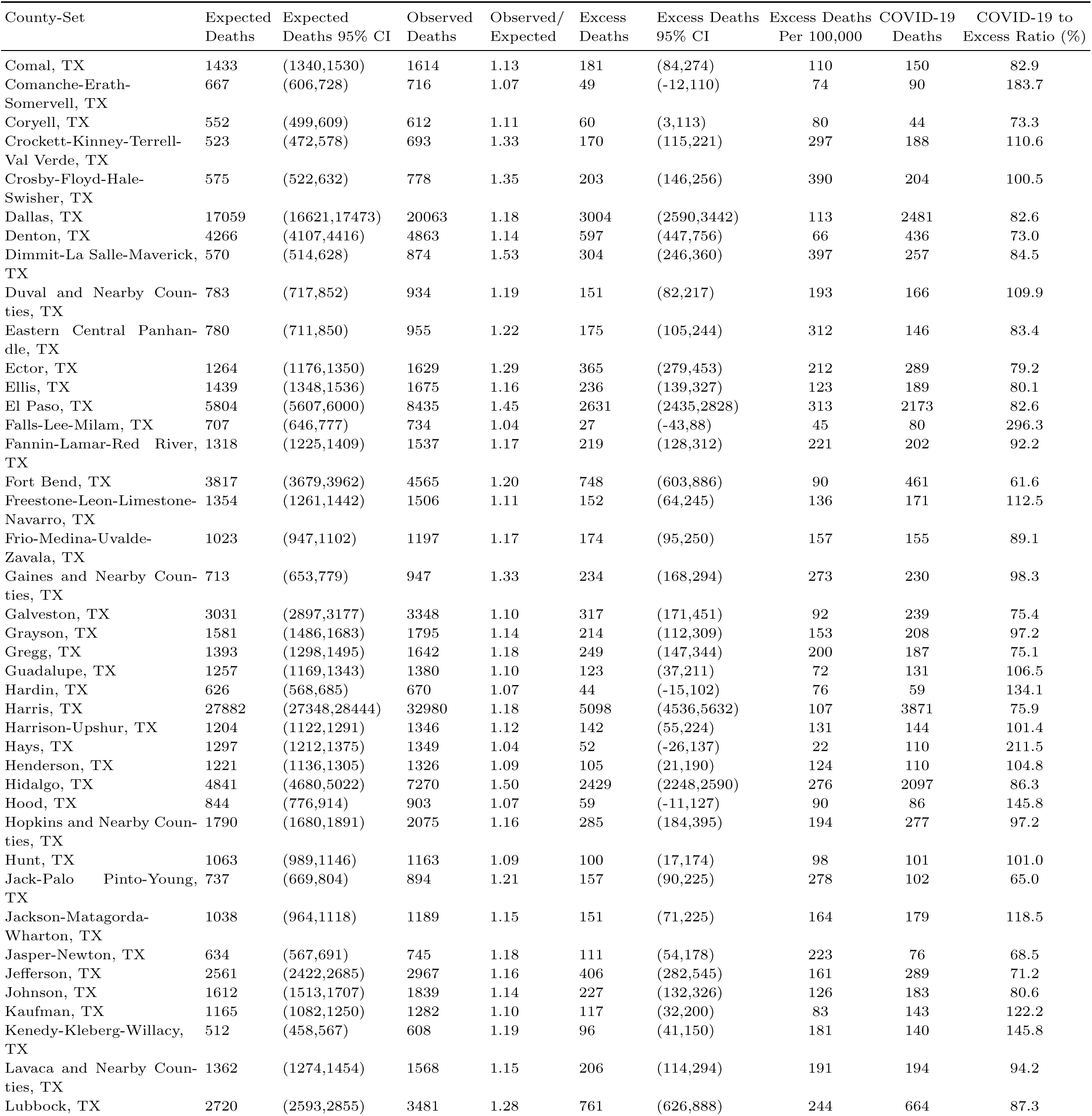
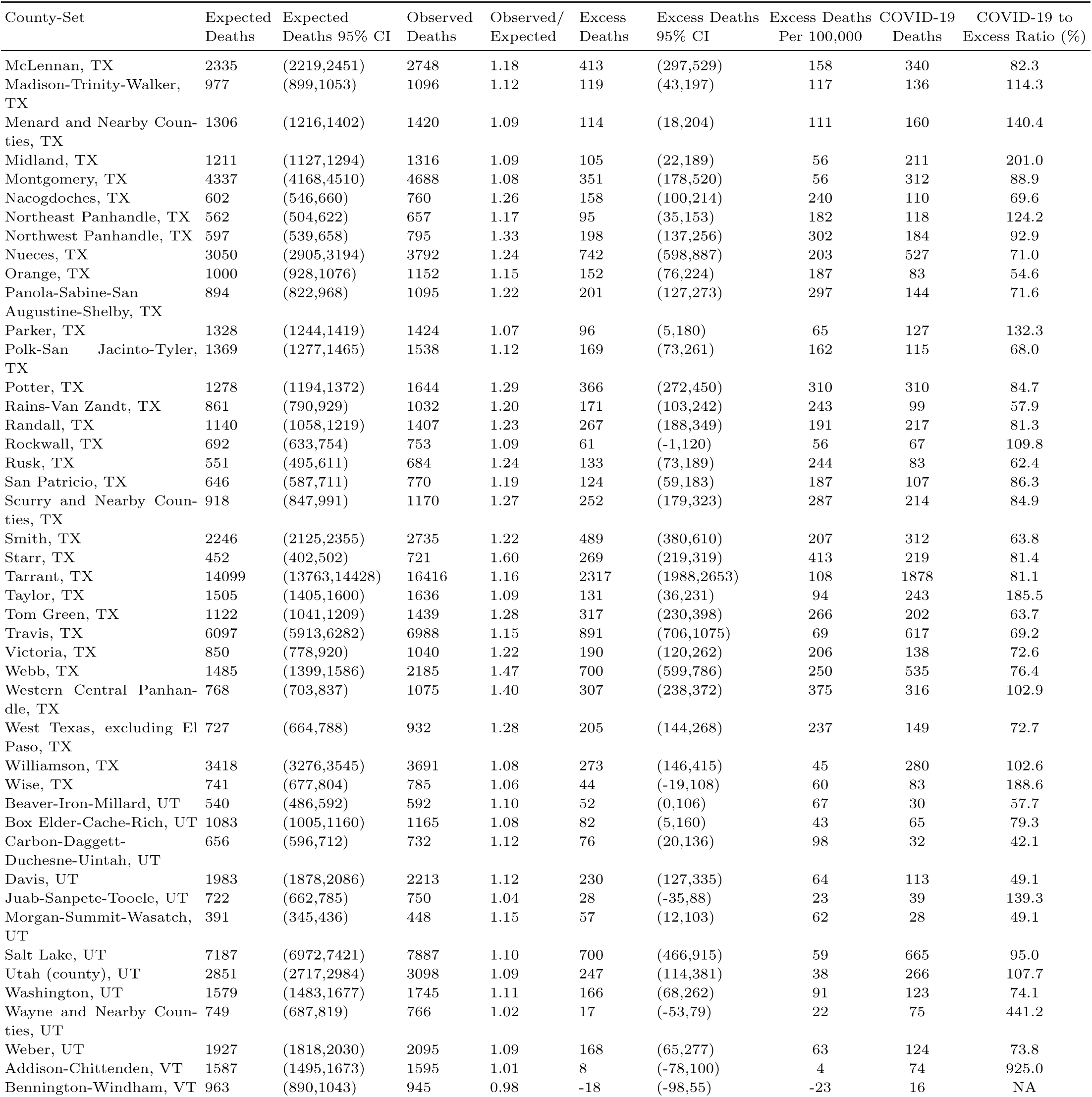
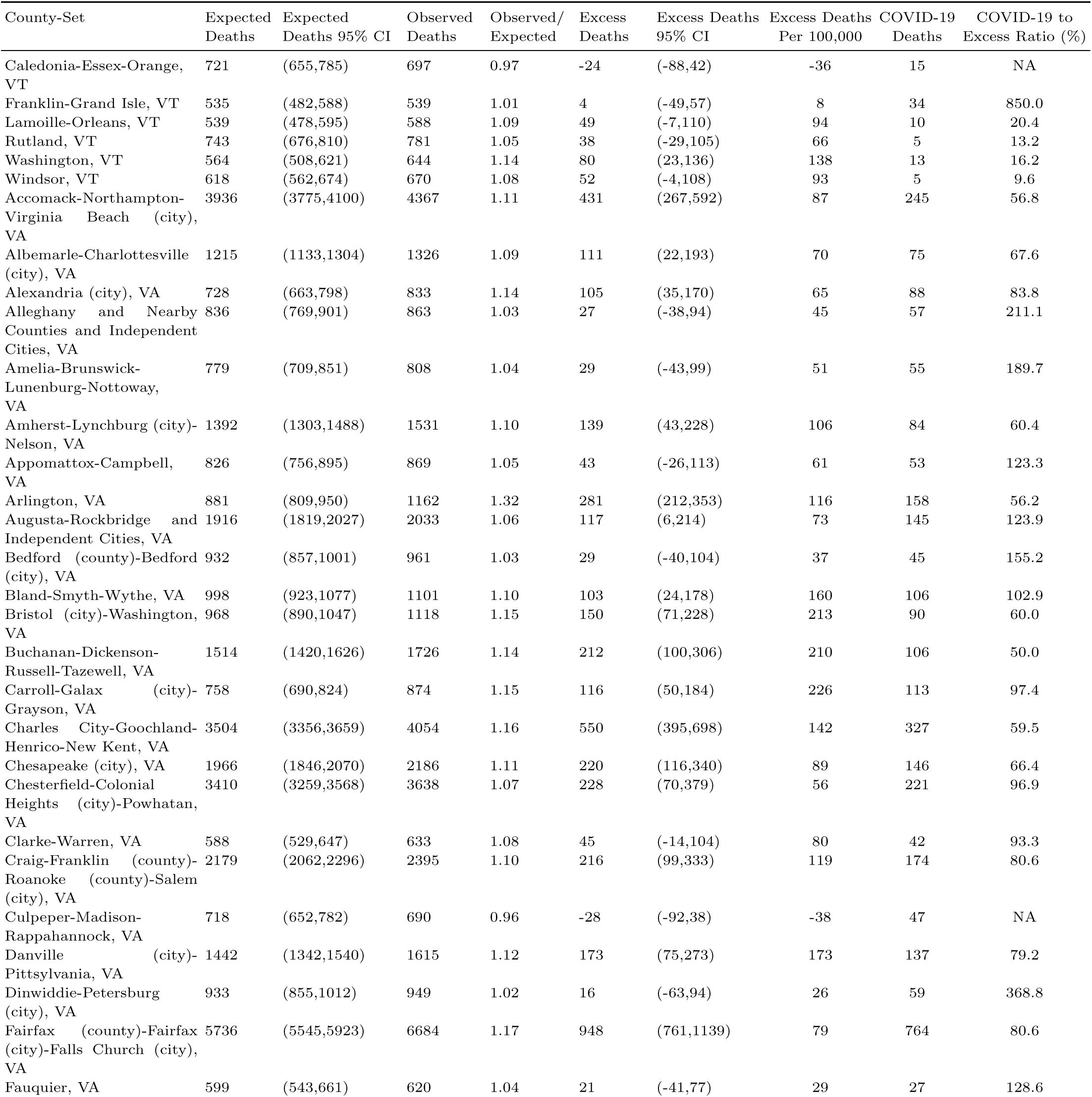
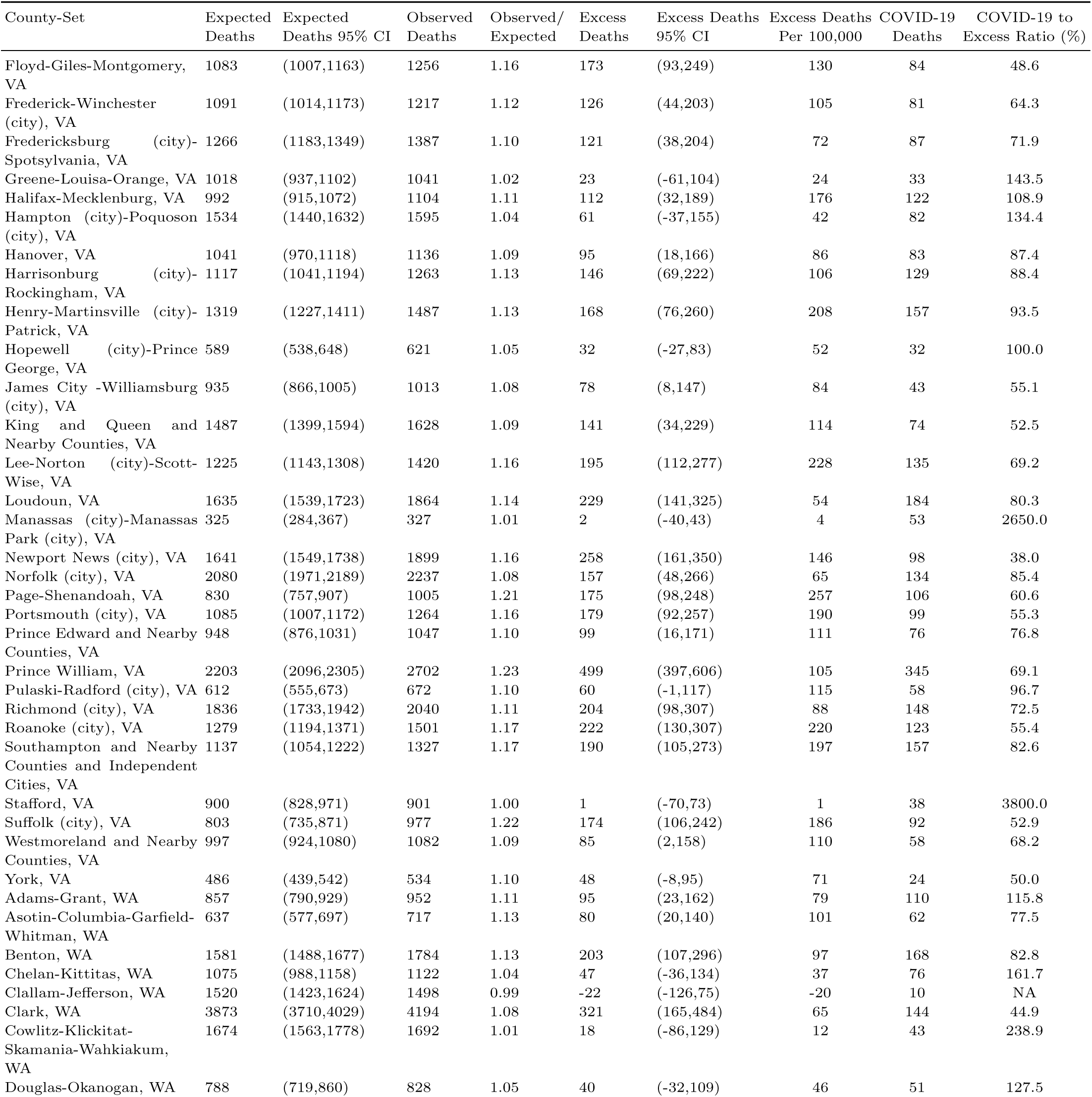
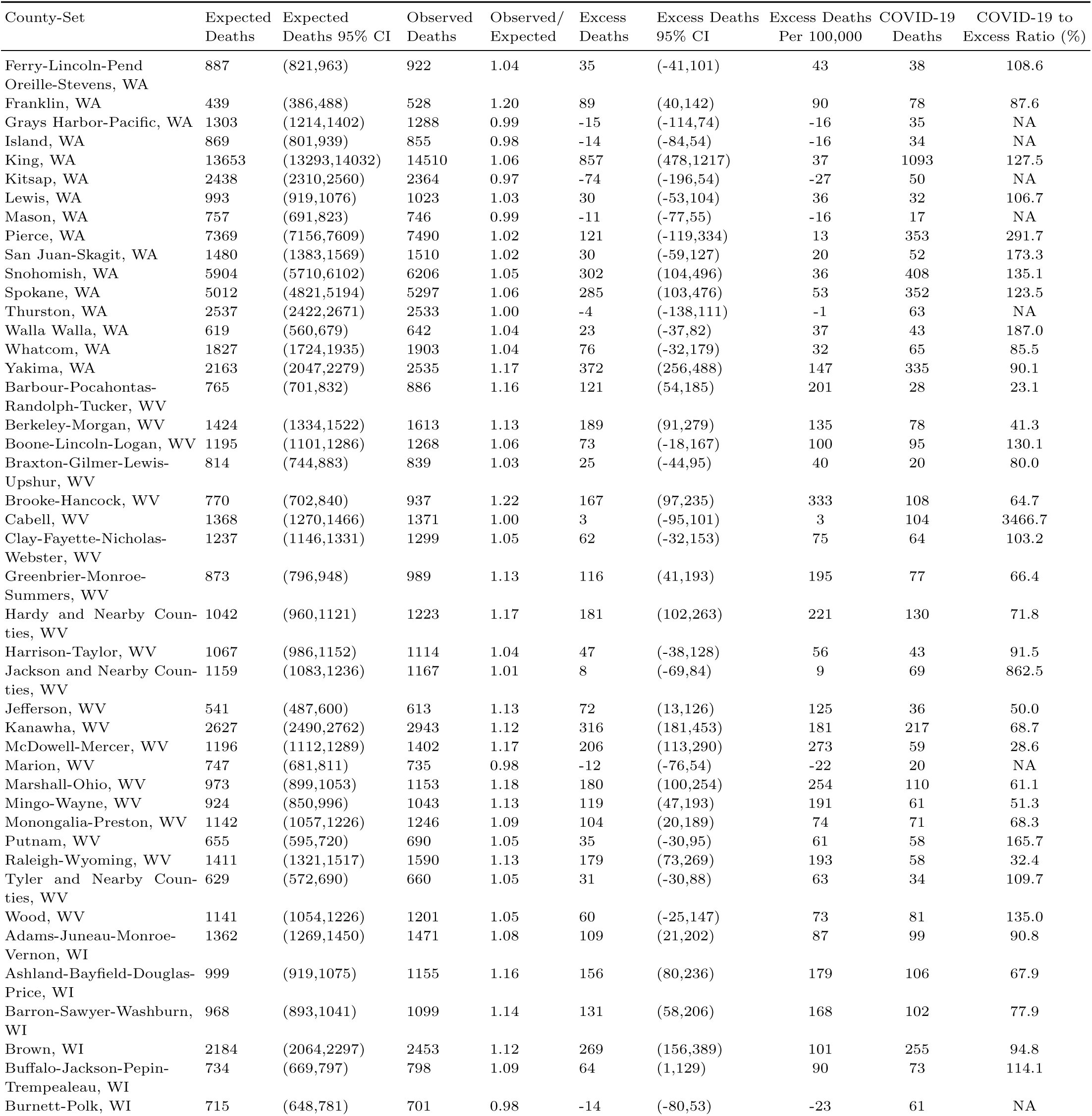
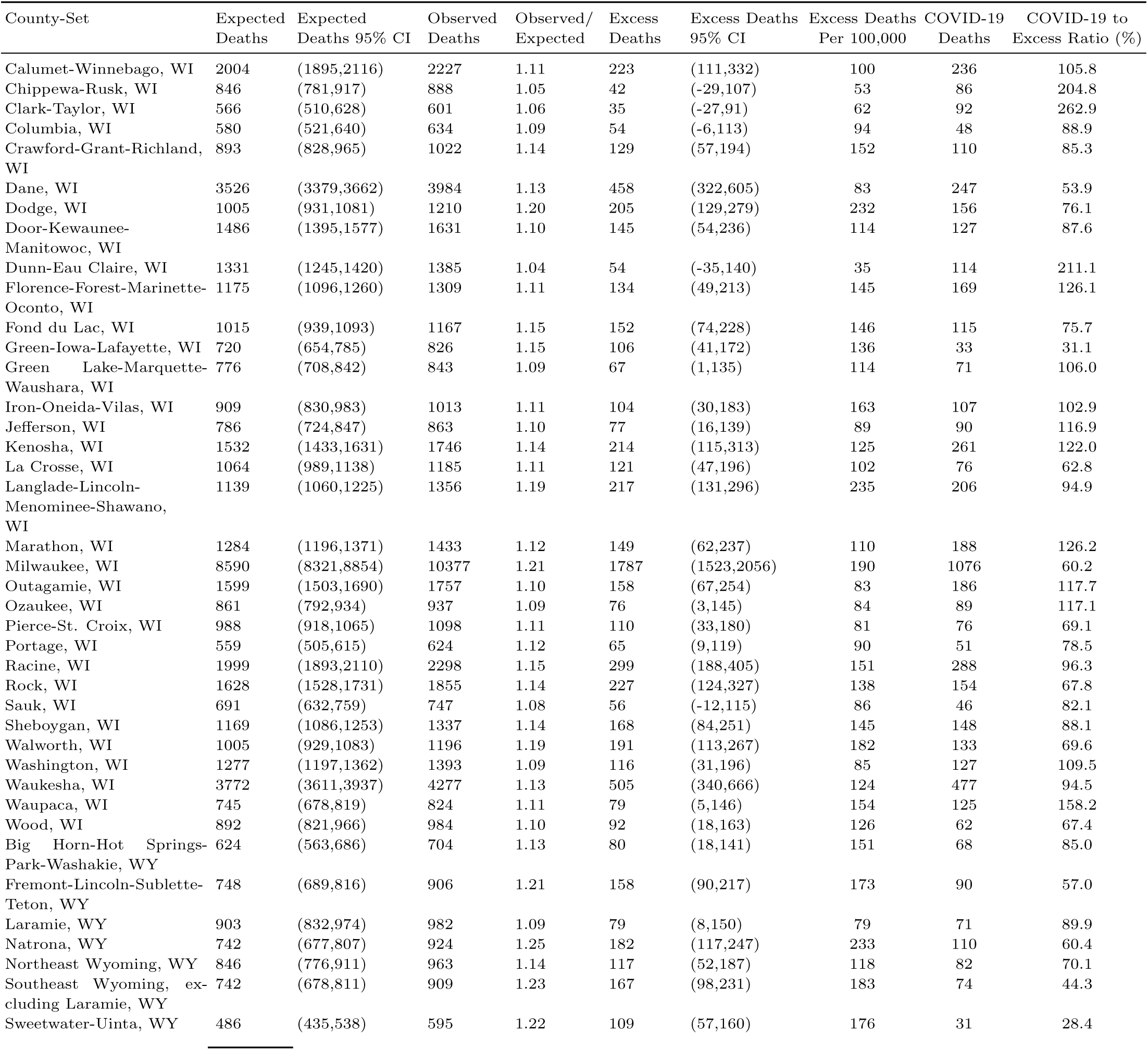
Primary Estimates for Each County-Set

## Footnotes

1 Data can be accessed at: https://data.cdc.gov/NCHS/AH-County-of-Residence-COVID-19-Deaths-Counts-2020/75vb-d79q

2 Detailed information on Census county-sets can be found at https://www.census.gov/geographies/reference-files/2000/demo/eeo/county-sets.html

3 The GitHub repository can be accessed at: https://github.com/pophealthdeterminantslab/county-level-estimates-of-excess-mortality

4 See, for example, McCullaugh and Nelder (1989)[20]

5 Details on how our exact specification was chosen are given in Appendix B

6 It may be desirable to explicitly include measures of such information to the extent that they vary significantly within counties over time. At present, we exclude such variables as few official county-level estimates of this type of information is presently available for 2020.

7 There could be positive serial correlation due persistent shocks, such as natural or other disasters, or there could be negative serial correlation due to survivorship bias.

8 We selected 2011 as the first year in our sample by comparing the predictive performance of our baseline Poisson GLM on 2019 mortality using different starting windows from 2009 to 2015.

9 The negative binomial model was infeasible to estimate with our high-dimensional fixed effects

10 Using alternative years to predict mortality results in the same general pattern of results, but with the Poisson and Gaussian specifications flip-flopping in terms of out-of-sample accuracy.

11 It is worth pointing out that, in addition to providing good fit and predictive performance, the Poisson model has attractive robustness properties. Most importantly, achieving consistent parameter estimates does not depend on the assumption of a Poisson distribution. Moreover, our model allows for overdispersion or underdispersion of the outcome variable and there is no restriction on time dependence within clusters. See, for example, Gourieroux, Monfort, and Trognon (1984)[34] and Wooldridge (1999)[35] for details.

12 In the standard Poisson GLM, the variance of the outcome variable is assumed to be equal to the mean. In practice this is highly restrictive, and so variance functions that permit overdispersion are common. Cameron and Trivedi (2013)[36] detail a number such specifications.

13 This procedure is based on Gelman and Hill (2006)[37]. Woolf et al. (2020) use a similar procedure in an analogous setting at the state level to generate predictive intervals.[38]

